# Early Genetic Evolution of Driver Mutations in Uveal Melanoma

**DOI:** 10.1101/2025.06.10.25329358

**Authors:** James J. Dollar, Christina L. Decatur, Ezekiel Weis, Amy C. Schefler, Miguel A. Materin, Timothy S. Fuller, Alison H. Skalet, David A. Reichstein, Ivana Kim, Kisha D. Piggott, Hakan Demirci, Thomas A. Aaberg, Prithvi Mruthyunjaya, Basil K. Williams, Eugene Shildkrot, Scott C.N. Oliver, Devron H. Char, Antonio Capone, John Mason, Scott D. Walter, Michael Altaweel, Jill Wells, Dan S. Gombos, Jay Duker, Peter Hovland, Tony Tsai, Cameron Javid, Michael A. Durante, Kyle R. Covington, Song Zhang, Zelia M. Correa, J. William Harbour

## Abstract

Uveal melanoma (UM) is an aggressive cancer of the eye that frequently results in metastatic death. UMs are most likely to metastasize when they are small, at a time when they are difficult to distinguish from benign nevi and often observed without treatment. Unfortunately, little is known about the early genetic evolution of UM or potential biomarkers to indicate small tumors undergoing malignant transformation. Here, we performed targeted next generation sequencing for the 7 canonical UM driver mutations in 1140 primary UMs, including 131 small early-stage tumors. We found that the evolutionary burst of genetic aberrations that determines the archetypal UM subtypes and metastatic propensity has already occurred by the time most small tumors are biopsied, although a significantly larger proportion of small tumors are still evolving compared to larger tumors. We found that the 15-gene expression profile (15-GEP) support vector machine discriminant score was the best indicator of tumors in transition from low-risk Class 1 to high-risk Class 2 signature. While *BAP1*, *SF3B1* and *EIF1AX* mutations were associated with poor, intermediate and good prognosis, respectively, mutation analysis was inferior to the prospectively validated 15-GEP + *PRAME* expression classifier for predicting metastasis-free and overall survival. These results provide a more complete picture of genetic evolution in UM, and they move us closer to a molecular definition of malignant transformation in this cancer type.

## INTRODUCTION

Uveal melanoma (UM) is a deadly cancer of the eye with a high propensity for metastasis^1^. UM can be divided into four prognostically significant subtypes based on a 15-gene expression profile (15-GEP; Class 1 or Class 2) combined with the expression status of the cancer-testis antigen *PRAME* (negative or positive)^2–6^. This 15-GEP/PRAME classifier was recently validated in the Collaborative Ocular Oncology Group Study Number 2 (COOG2), a large international prospective multicenter biomarker study^6^. Within this molecular landscape, there are two clusters of highly recurrent UM-associated mutations (UMAMs)^7^. The first cluster consists of initiating mutations in one of four members of the G_q_ signaling pathway (*GNAQ*, *GNA11*, *CYSLTR2* and *PLCB4*)^8–12^. G_q_ mutations do not appear to be sufficient for malignant transformation without the acquisition of further genomic aberrations, as they are also found in benign uveal nevi^8,9,13–15^. The second cluster comprises the “BSE” (*BAP1*, *SF3B1* or *EIF1AX*) mutations which are thought to signify malignant transformation and are associated with high, intermediate and low metastatic risk, respectively^16–19^. Mutations in *GNAQ*, *GNA11*, *CYSLTR2*, *PLCB4*, *SF3B1*, and *EIF1AX* are small somatic variants that are easily detected by next generation sequencing (NGS). In contrast, mutations in *BAP1* comprise a variety of deleterious alterations, some of which can be challenging to detect with NGS^20^. *BAP1* mutations are usually somatic but occasionally arise in the germline, and they become fully manifest by loss of the other allele by whole chromosome 3 loss^16,21^. BSE mutations and associated copy number variations (CNVs) arise in the primary tumor around the same time during a punctuated evolutionary burst^14,20,22,23^, although the timing of this event during genetic evolution remains unclear.

Since UM is thought to micrometastasize early, when tumors are small ^24^, thereby explaining the high metastatic rate despite successful primary tumor treatment ^25^, there remains a critical unmet need to elucidate the early genetic events in UM tumorigenesis and to better understand the molecular transition from benign nevus to malignant melanoma. Unfortunately, our current understanding of UM genetic evolution is inferred almost exclusively from large primary tumors that were treated by enucleation (eye removal)^14,20,22,23^ or from metastatic tumors ^26,27^. This lack of knowledge regarding small tumors is due in large part to their being treated by observation or eye-sparing therapies where genetic analysis is limited to small biopsy samples. We developed and analytically validated a targeted NGS panel for robust detection of all 7 recurrent UMAMs using residual tumor biopsy material obtained during standard of care prognostic testing^28^. Here, we characterize the mutation landscape, infer early genetic evolution, and evaluate the prognostic significance of UMAMs in the largest multicenter prospective study to date in a real-world cohort of cases across the full spectrum of UM tumor size.

## RESULTS

### Patient cohort

Of 1687 subjects enrolled in COOG2, 1140 met inclusion criteria for this report, which included the presence of at least one UMAM (**Supplementary Fig. 1**). Baseline demographic and clinical information are summarized in **Supplementary Table 1**. Median age at study entry of 64.3 years (range, 18–99 years), including 550 (48.3%) female patients and 590 (51.8%) male patients. Baseline tumor thickness averaged at 5.5 mm (ranging 1.0 to 18.0 mm), while the mean tumor diameter was 12.6 mm (ranging 3.0 to 28.9 mm). Ciliary body involvement was present in 201 (17.6%) of the tumors. 15-GEP was Class 1 in 716 (62.8%) cases and Class 2 in 424 (37.2%) cases. *PRAME* expression was negative in 757 (66.4%) cases and positive in 383 (33.6%) cases. Median follow-up was 52.8 months. Metastatic disease was detected in 229 (20.1%) patients, and the median time to metastasis among patients with an event was 21.9 months (range, 17.3–79.9 months). Local tumor recurrence was identified in 54 (4.7%) patients with a median time of 28.5 months (range, 3.5–82.2 months) after biopsy/primary enucleation, with 28 (51.9%) of these patients subsequently developing metastatic disease.

### Landscape of uveal melanoma associated mutations

UMAM NGS results are summarized in **Fig. 1a-c** and **Supplementary Table 2** and **Supplementary Fig. 2**. G_q_ mutations were detected in *GNAQ* in 558 (48.9%), *GNA11* in 530 (46.5%), *PLCB4* in 25 (2.2%), and *CYSLTR2* in 14 (1.2%) cases. BSE mutations were detected in *BAP1* in 364 (31.9%), *SF3B1* in 194 (17.0%), and *EIF1AX* in 304 (26.7%) cases. Associations between UMAMs and clinical and molecular features are summarized in **Supplementary Table 3**. *GNAQ* mutations were associated with Class 1 tumors (*P*<0.0001), decreased patient age (*P*=0.008), decreased tumor diameter (*P*=0.003) and tumor thickness (*P*=0.0006). Conversely, *GNA11* mutations were associated with Class 2 tumors (*P*=0.0005), *PRAME*(+) status (*P*=0.05), increased patient age (*P*=0.002), increased tumor diameter (*P*=0.004) and tumor thickness (*P*=0.002), and they showed near-significant association with ciliary body involvement (*P*=0.06). G_q_ mutations were mutually exclusive, except for 6 (0.5%) cases in which a *GNAQ*^Q209P^, *GNA11*^Q209L^, *GNAQ*^R183Q^ or *GNA11*^R183C^ recurrent hotspot mutation was accompanied by a rare *GNAQ*^P193T^, *GNAQ*^T175M^, *CYSLTR2*^S154N^ or *PLCB4*^D630N^ mutation (**Fig. 1d**). *BAP1* mutations were associated with Class 2 tumors (*P*<0.0001), *PRAME*(+) status (*P*<0.0001), increased patient age (*P*<0.0001), increased tumor diameter (*P*<0.0001), increased tumor thickness (*P*<0.0001), ciliary body involvement (*P*<0.0001), mutations in *GNA11* (*P*=0.01), *PLCB4* (*P*=0.02) and *CYSLTR2* (*P*=0.02), and absence of mutations in *GNAQ* (*P*<0.0001). The spectrum of *BAP1* mutation types did not differ significantly between Class 1 and Class 2 tumors (**Fig. 1c**). *SF3B1* mutations were associated with Class 1 tumors (*P*<0.0001), *PRAME*(+) status (*P*<0.0001), decreased patient age (*P*<0.0001), increased tumor diameter (p=0.02), and brown iris color (*P*=0.008). *EIF1AX* mutations were associated with Class 1 tumors (*P*<0.0001), PRAME(-) status (*P*<0.0001), decreased tumor diameter (*P*<0.0001), lack of ciliary body involvement (*P*<0.0001), and male sex (*P*<0.0001). By and large, BSE mutations were mutually exclusive, with only 26 (2.3%) cases harboring two BSE mutations, including *BAP1* and *SF3B1* in 5 cases, *BAP1* and *EIF1AX* in 15 cases, and *SF3B1* and *EIF1AX* in 6 cases (**Fig. 1d**). No cases harbored all three BSE mutations.

**Fig. 1.**
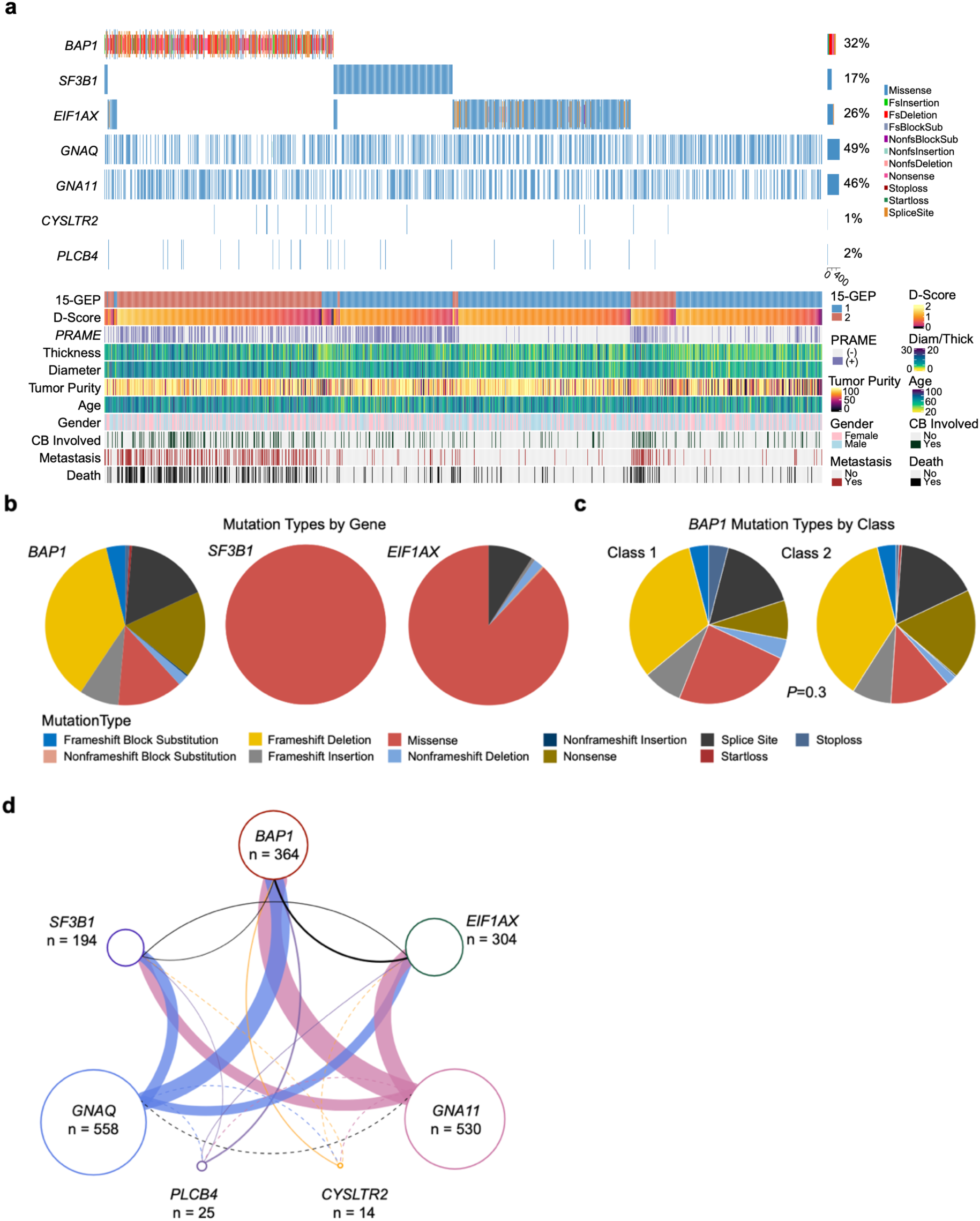
Genetic landscape of uveal melanomas. **a**, Oncoprint of 1140 primary uveal melanomas, demonstrating the 7 canonical uveal melanoma associated mutations (UMAMs), along with 15-GEP status, PRAME status, tumor thickness (millimeters), tumor diameter (millimeters), gender, metastatic status (yes or no), and survival status (alive or dead). **b-c,** Pie charts summarizing variant types for *BAP1*, *SF3B1*, and *EIF1AX* mutations **b,** for all samples with at least one mutation (n=836) and **c**, for *BAP1* mutations in Class 1 (n=25) and Class 2 tumors (n=339). Significance was calculated by two-tailed Fisher’s exact test. **d,** Connectivity plot indicating co-occurring mutations, with connector color representing G_q_ mutation (blue, *GNAQ*; mauve, *GNA11*; purple, *PLCB4*; yellow, *CYSLTR2*), and connector thickness corresponding to the number of cases. Dashed lines indicate < 2 cases. Diam, tumor diameter; Thick, tumor thickness. CB, ciliary body. D-score, 15-GEP support vector machine discriminant score. Variant types described in Materials and Methods.

### Prognostic significance of UMAMs

The prognostic significance of each UMAM was evaluated by Cox regression. In univariate analysis (**Supplementary Table 4**), *BAP1* was the only UMAM associated with shorter MFS (HR=5.9, *P<*0.0001), whereas *EIF1AX* (HR=0.2, *P<*0.0001), *SF3B1* (HR=0.5, *P=*0.0009), and *GNAQ* (HR=0.8, *P=*0.05) mutations were associated with longer MFS (**Supplementary Fig. 3**). *BAP1* (HR=4.3, *P<*0.0001) and *GNA11* (HR=1.4, *P*=0.007) mutations were associated with shorter OS, whereas *EIF1AX* (HR=0.4, *P<*0.0001), *SF3B1* (HR=0.5, *P=*0.0005), and *GNAQ* (HR=0.7, *P=*0.001) were associated with longer OS (**Supplementary Fig. 4**). In multivariate Cox analysis of MFS (**Supplementary Table 5**), when 15-GEP was entered into the model, mutations in *BAP1*, *EIF1AX*, *GNAQ*, and *GNA11* were rendered non-significant, and mutations in *SF3B1* became associated with shorter (rather than longer) MFS (HR=1.7, *P*=0.03). In multivariate Cox analysis of OS, all UMAMs became non-significant when 15-GEP was entered into the model. Among Class 1 tumors, when *PRAME* status was entered into a multivariate Cox model, mutations in *SF3B1* became non-significant for MFS and OS. Thus, the combination of 15-GEP and *PRAME* renders all UMAMs non-significant and redundant for prognostic testing in UM.

### Insights into early genetic evolution from small tumors

To date, almost all genetic studies in UM have been performed on large enucleated tumors^20,22^, but these represent only a small minority of the most advanced cases^1,29^. We hypothesized that smaller tumors, which are usually treated with eye-sparing therapies or observed for growth prior to treatment ^30^, may reveal new insights into the early genetic evolution of UM. Thus, we compared 131 small tumors (defined as having thickness ≤ 2.5 mm and diameter ≤ 12mm based on thresholds established in previous reports using the 15-GEP^30,31^ to the remaining 1009 larger tumors (**Fig. 2a-c and Supplementary Table 6**). Small tumors were more likely than larger tumors to be Class 1 (*P*<0.0001), *PRAME*(-) (*P*=0.004), *BAP1*^wt^ (*P*=0.0006), and to lack any BSE mutation (*P*=0.001), suggesting that most or all UM begin as small Class 1 tumors that later acquire a BSE mutation during tumor growth. Average tumor purity was lower for small tumors (mean, 58.6% ± 3.1%) compared to larger tumors (mean, 81.9% ± 0.8%)(Wilcoxin test, *P*<0.0001). Additionally, the discriminant score – the distance a given sample from the 15-GEP SVM decision boundary^32^ – was significantly lower in small versus larger Class 2 tumors (*P*=0.003)(**Fig. 2d**), potentially suggesting that small Class 2 tumors with low discriminant scores may have recently transitioned from small Class 1 tumors.

**Fig. 2.**
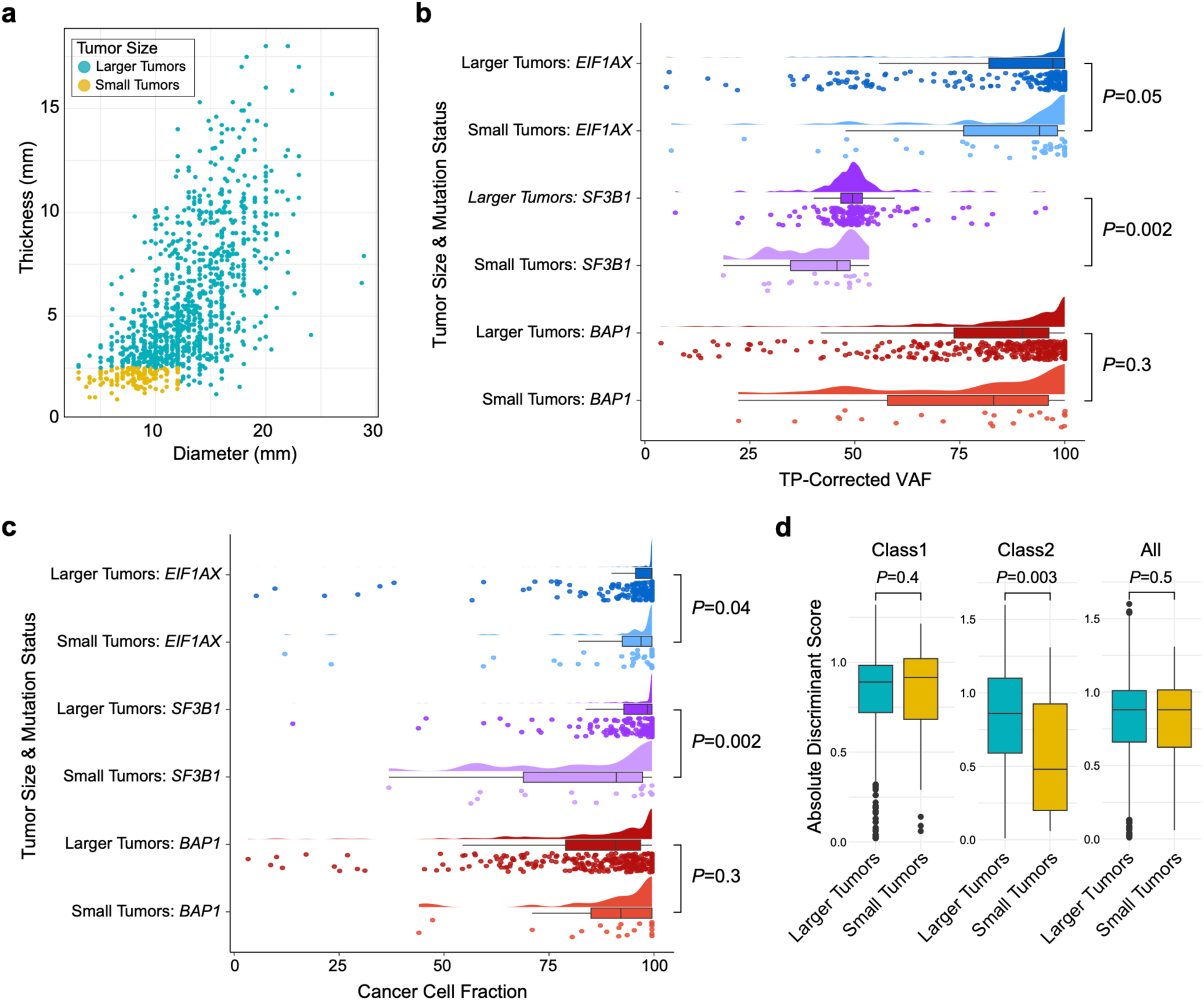
Comparison of cancer cell fraction and 15-GEP discriminant score in small versus larger uveal melanomas. **a**, Scatter plot displaying the distribution of tumor thickness and diameter for 131 small tumors (yellow dots) versus 1009 larger tumors (green dots). **b,** Raincloud plot of TP-corrected VAF for each BSE mutation in small versus larger tumors. **c,** Raincloud plot of cancer cell fraction (CCF) for each BSE mutation in small versus larger tumors. **d,** Box plot comparing the 15-GEP discriminant score for small tumors (yellow boxes) versus larger tumors (green boxes), comparing Class 1, Class 2 and all tumors. Continuous variables were compared by two-tailed Wilcoxon rank-sum test. BSE, mutation in *BAP1*, *SF3B1* or *EIF1AX*; CCF_BSE_, cancer cell fraction for each BSE mutation; TP, tumor purity; VAF, variant allele frequency; mm, millimeter.

### Insights into early genetic evolution from discordant tumors

While most cases exhibited the expected relationships between 15-GEP Class and *BAP1* status, there was a small subset of discordant cases (**Fig. 3a-f**), including 25 (3.5%) Class 1 tumors with a *BAP1* mutation (**Fig. 4a-b** and **Supplementary Table 7**). While *BAP1* mutation types did not differ significantly between Class 1 and Class 2 tumors (**Fig. 1c**), we further investigated potential functional differences in *BAP1* mutations between the two tumor classes using the recently described saturation genome editing (SGE) database for BAP1 ^33^. After excluding 106 complex *BAP1* mutations involving ≥5 nucleotide alterations, we successfully mapped 218 of the remaining 258 (>80%) *BAP1* mutations to the database. 97.7% of *BAP1* mutations (213/218) were functional classified as “depleted”, indicating a high concordance between our mutation-calling methodology and the SDE methodology (**Supplementary Fig. 5a**). Importantly, there was no significant difference in deleterious categorization (*P*=0.3) or functional scores (*P*=0.3) for *BAP1* mutations in Class 1 versus Class 2 tumors (**Supplementary Fig. 5a-b**).

**Fig. 3.**
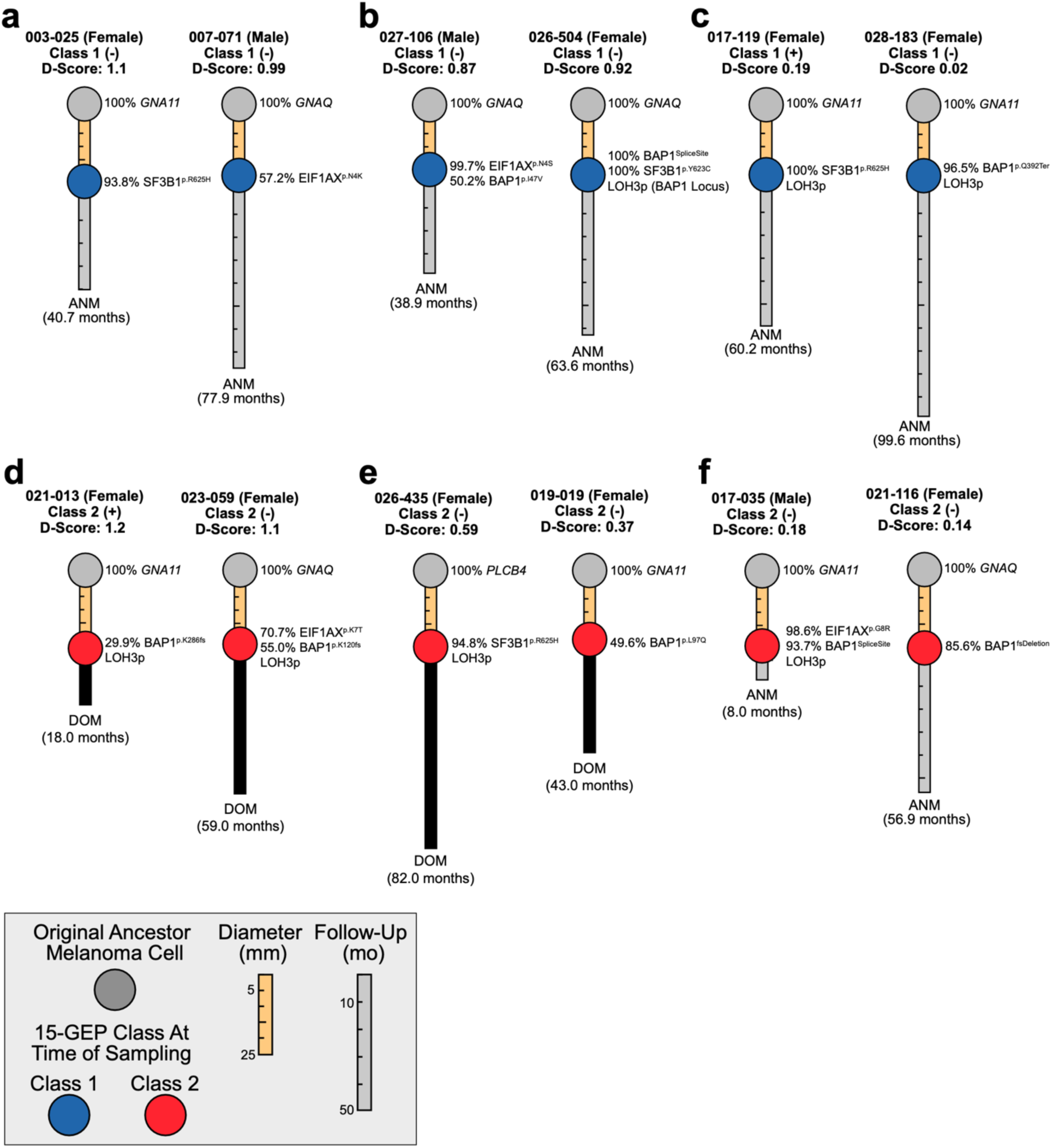
Insights into early genetic evolution from 15-GEP discriminant score and BSE cancer cell fraction. **a**, Typical Class 1 tumors with high discriminant scores: Case #003-025, near-clonal *SF3B1* mutation; Case #007-071, sub-clonal *EIF1AX* mutation. **b,** Discordant Class 1 tumors with high discriminant scores: Case #027-106, near-clonal *EIF1AX* mutation and subclonal *BAP1* mutation; Case #026-504, clonal *BAP1* and *SF3B1* mutations and partial LOH3p involving a limited region around the *BAP1* locus. **c,** Discordant Class 1 tumors with low discriminant scores: Case #017-119, clonal *SF3B1* mutation and LOH3p; Case #028-183, near-clonal BAP1 mutation, LOH3p and very low discriminant score (0.02). **d,** Class 2 tumors with high discriminant scores and bi-allelic *BAP1* loss: Case #021-013, sub-clonal *BAP1* mutation; Case 023-059, near-clonal *EIF1AX* mutation and subclonal *BAP1* mutation. **e,** Class 2 tumors with intermediate discriminant scores: Case #026-435, near-clonal *SF3B1* mutation and LOH3p but no detectable *BAP1* mutation; Case #019-019, sub-clonal *BAP1* mutation and no detectable LOH3p. **f,** Class 2 tumors with low discriminant scores: Case #017-035, near-clonal *EIF1AX* and *BAP1* mutations with LOH3p; Case #021-116, near-clonal BAP1 mutation with no detectable LOH3p. The length of the connector between the ancestor melanoma cell and the melanoma is proportional to tumor diameter (in millimeters) at the time of tumor sampling. The length of the extension beyond the melanoma is proportional to time to death or last follow-up (in months) with final status indicated. ANM, alive no metastasis; DOM, dead of metastasis; D-score, support vector machine discriminant score; LOH3p, loss of heterozygosity of chromosome 3; (-), *PRAME* negative; (+), *PRAME* positive. Mutation nomenclature described in Material and Methods.

**Fig. 4.**
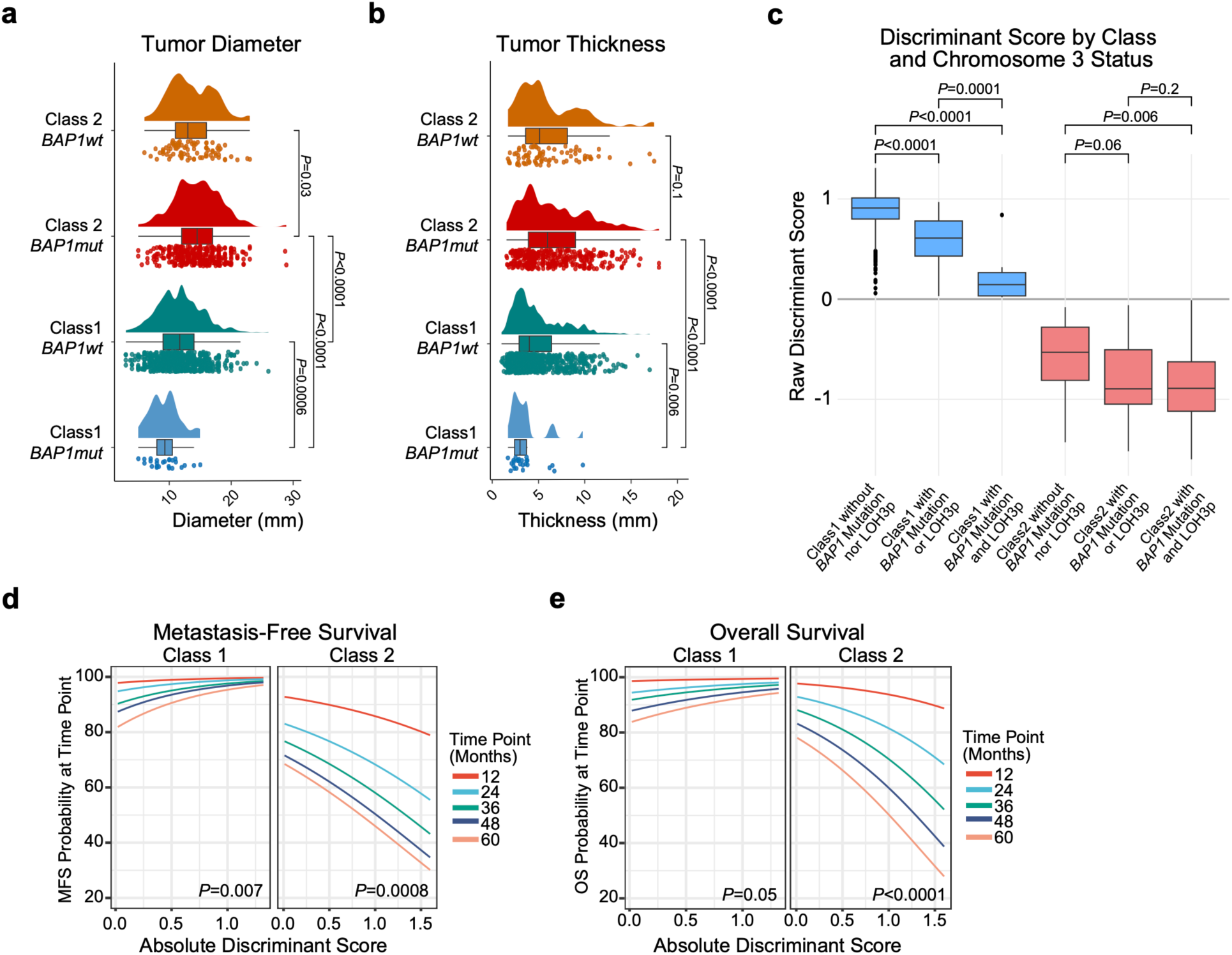
Features of *BAP1* mutations by 15-GEP Class status. **a**, Raincloud plot depicting tumor diameter in relation to 15-GEP Class and *BAP1* mutation status (n=1140). **b,** Raincloud plot depicting tumor thickness in relation to 15-GEP Class and *BAP1* mutation status (n=1140)**. c**, Box plot comparing raw discriminant scores by 15-GEP Class and *BAP1* allelic “dosage” reflected in *BAP1* mutation and LOH3p status (n=905). **d-e**, Survival analysis plots displaying the **d**, metastasis-free survival and **e**, overall survival probabilities for Class 1 (n=715) and Class 2 (n=418) UM according to absolute discriminant score at specified time points ranging from 12 to 60 months. Significance for continuous variables was determined by two-tailed Wilcoxon rank-sum test. Significance for survival analysis was calculated by Cox proportional hazard analysis.

Class 1/*BAP1*^mut^ tumors were associated with decreased tumor diameter (*P*=0.0006), decreased tumor thickness (*P*=0.006), and decreased discriminant score (*P*<0.0001) compared to Class 1/*BAP1*^wt^ tumors, suggesting that (1) most Class 1 tumors that acquire *BAP1* mutations do so when they are small and then convert to Class 2 before they have grown to a larger size, (2) *BAP1* mutations may be less likely to arise in Class 1 tumors above a certain size, possibly because the selective advantage has been satisfied by another aberration (e.g., *SF3B1* or *EIF1AX* mutation), and (3) the transition from Class 1 to Class 2 after acquiring a *BAP1* mutation is accompanied by a progressive decrease in the discriminant score on the Class 1 side of the decision boundary before increasing on the Class 2 side. Further, there was no difference in MFS or OS between Class 1/*BAP1*^mut^ tumors compared to all Class 1/*BAP1*^wt^ tumors (**Supplementary Fig. 3-4**), nor compared to a propensity score matched cohort of 75 Class 1/*BAP1*^wt^ tumors (**Supplementary Fig. 6a-c**). The lack of survival difference could be explained by several factors: (1) since Class 1/*BAP1*^mut^ tumors are generally small, any real decrease in survival may be very small and require longer follow-up to be detected, and (2) patients with Class 1/*BAP1*^mut^ tumors may be among those most likely to be “cured” by effective local treatment by preventing early micrometastasis. Longer follow-up will be required to discern between these possibilities.

### Insights into early genetic evolution from discriminant score and cancer cell fraction

Since the Class 2 signature results from bi-allelic loss of *BAP1*^16^, we inferred the temporal relationship between *BAP1* loss and 15-GEP switch from Class 1 to Class 2 using the SVM discriminant score and cancer cell fraction for *BAP1* (CCF*_BAP1_*) in a subgroup of 905 cases in which copy number status was available for the *BAP1* locus at chromosome 3p21. As anticipated, progressive decrease in BAP1 protein “dosage” (via mutational inactivation or chromosomal loss of the gene) was accompanied by a shift from Class 1 to Class 2 and an inversion of the discriminant score (**Fig. 4c**), with lower discriminant scores being associated with worse outcome in Class 1 tumors and better outcome in Class 2 tumors (**Fig. 4d-e**). We next evaluated cancer cell fractions for each BSE mutation. As expected, increasing CCF*_SF3B1_* and CCF*_EIF1AX_* were associated with larger tumor size (**Supplementary Table 8**), suggesting that these mutations usually arise in small tumors and progressively outcompete preexisting UM cells during tumor growth. Unexpectedly, however, there was no association between CCF_BAP1_ and tumor size (**Supplementary Table 8**), nor was there an association between CCF*_BAP1_* and discriminant score (Spearman correlation, R = −0.1, *P*=0.07), MFS or OS (**Supplementary Table 9**). Taken together, these findings suggest that BSE mutations usually occur early in the genetic evolution of UM when tumors are small. However, in the case of *BAP1*, mutational inactivation triggers a progressive transcriptomic shift from Class 1 to Class 2 accompanied by a decrease in discriminant score on the Class 1 side of the SVM decision boundary followed by an increase on the Class 2 side that is not tightly linked to CCF*_BAP1_* but likely also depends on alterations that *BAP1* loss causes to the tumor immune microenvironment (**Fig. 5**).

**Fig. 5.**
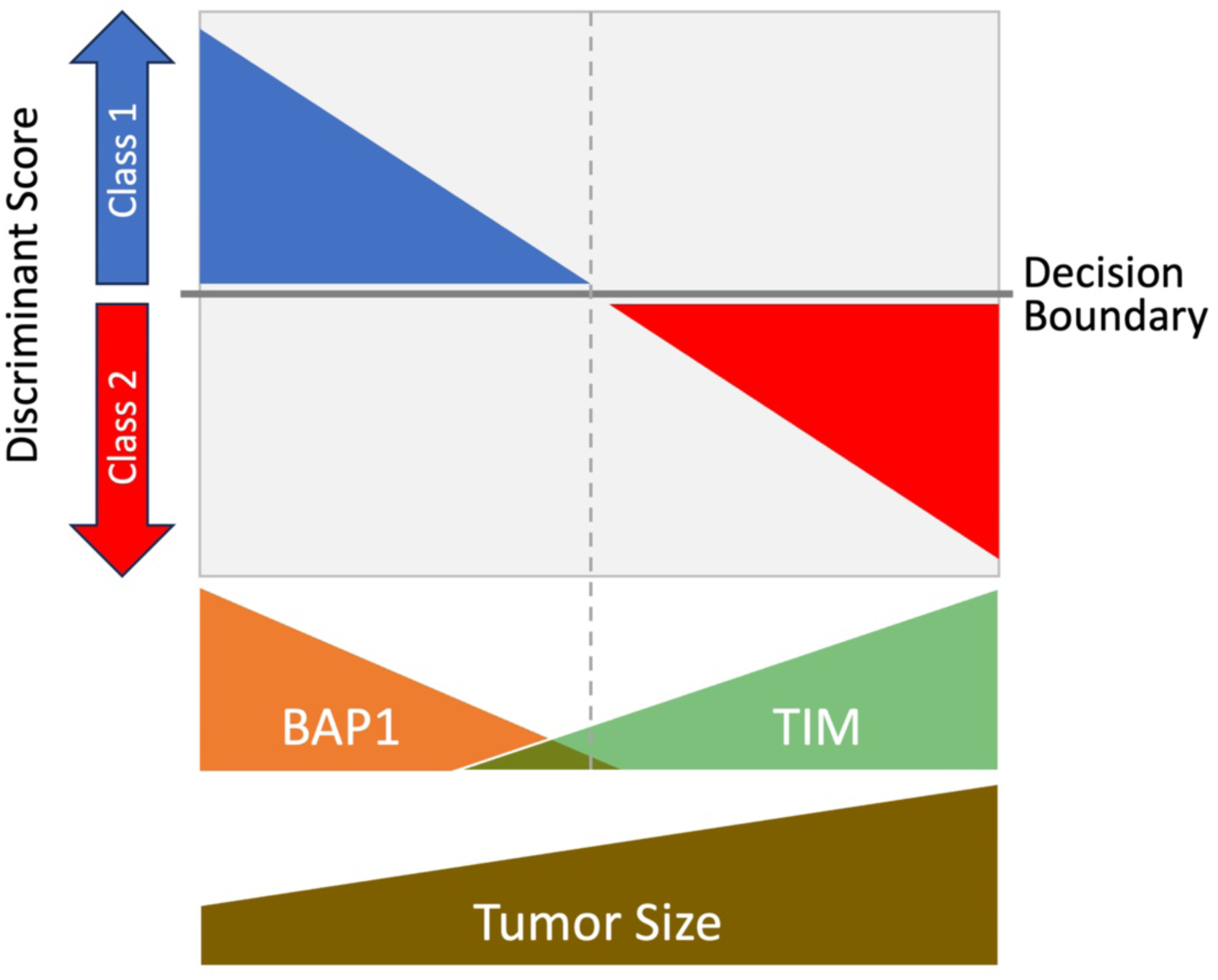
Hypothesis for relationship between *BAP1* dosage, tumor immune microenvironment, and discriminant score. *BAP1* dosage decreases as BAP1-deficient tumor cells outcompete BAP1-wildtype tumor cells, leading to altered composition of infiltrating immune cells in the tumor immune microenvironment (TIM). Since the 15-GEP includes genes expressed in tumor cells, immune cells or both, inversion of the SVM discriminant score from the Class 1 side to the Class 2 side of decision boundary occurs progressively as the transcriptional effects of *BAP1* loss accrue in both tumor and immune cells. This would explain why there is not a strict association between the fraction of cancer cells harboring mutant *BAP1* (CCF_BAP1_) and the discriminant score, as the rate at which the TIM changes following BAP1 loss may differ between individuals. This would also explain why transitional tumors with low discriminant score tend to be small, whereas larger tumors which have had longer for these transcriptional changes to occur tend to have high discriminant scores.

## DISCUSSION

This study provides the most comprehensive overview to date of the genetic landscape of UM across a real-world spectrum of tumor size, including small tumors previously excluded from most genetic analyses. Our findings suggest that the recurrent genomic aberrations that give rise to the archetypal evolutionary trajectories in UM^20^ usually arise early when tumors are small. We confirmed the high, intermediate and low metastatic risk associated with *BAP1*, *SF3B1* and *EIF1AX* mutations, respectively, but we found that these mutations are inferior to the 15-GEP/PRAME classifier for predicting metastasis-free and overall survival. Further, we found that the 15-GEP SVM discriminant score is a better indicator of tumors in transition from Class 1 to Class 2 than is the fraction of cancer cells harboring a *BAP1* mutation (CCF*_BAP1_*).

These findings provide new insights into the early genetic evolution of UM, explain the high metastatic rate despite successful primary tumor treatment, and allow further refinement of prognostic tools to guide the precision treatment of selected small tumors at an earlier stage that may improve survival.

An unresolved question in the field is whether Class 2 tumors arise from Class 1 tumors or from a distinct precursor cell. We found that small tumors were much more likely to be Class 1 and to lack *BAP1* mutations compared to larger tumors. Further, we identified tumors that appeared to be in transition between Class 1 and Class 2 with low discriminant score and/or subclonal *BAP1* mutations (**Fig. 3**), consistent with the hypothesis that most or all Class 2 tumors arise from Class 1 tumors following bi-allelic loss of *BAP1*. We confirmed that BSE mutations are usually mutually exclusive, but there were some cases in which an *SF3B1* or *EIF1AX* mutation was followed by a *BAP1* mutation, indicating that the former does not absolutely “protect” from the latter and that BSE mutations may occasionally coexist and compete during early tumor evolution. Small tumors were also more likely to lack any BSE mutation, suggesting that they were sampled early during genetic evolution and may not be fully transformed. The lower tumor purity in small lesions was likely due to an increased chance of aspirating surrounding normal cells and may have contributed to the lower detection rate of BSE mutations. However, since a Gq mutation was detected in all of these cases, a putative undetected BSE mutation would have necessarily been present at a low CCF, which is still consistent with our conclusion that some small tumors are still evolving their canonical UMAMs at the time of biopsy. It will be critical to determine how to distinguish between small tumors that can be safely monitored without treatment versus those that are likely to acquire high-risk genetic features if left untreated.

Among the 7 recurrent UMAMs, only the BSE mutations exhibited independent prognostic significance, and even the BSE mutations were insignificant when the 15-GEP/PRAME classifier was included in multivariate analysis for both metastasis free and overall survival. The superiority of gene expression profiling over mutational analysis may have several explanations. First, it is likely that some Class 2/*BAP1*^wt^ tumors actually had *BAP1* mutations that were undetectable with standard NGS methods^20^. Long read sequencing methods may improve the ability to detect such *BAP1* mutations^34^. Second, there was no significant correlation between CCF_BAP1_ and survival, and there was no CCF*_BAP1_* threshold where the 15-GEP switched from Class 1 to Class 2, suggesting that there are additional factors beyond CCF_BAP1_ that determine the Class 2 signature. Indeed, we previously showed that gene expression from tumor-infiltrating immune cells contributes substantially to the 15-GEP^23^ and that *BAP1* loss in UM cells alters gene expression in adjacent immune cells^35^. Consequently, the 15-GEP appears to represent a functional snapshot of the transcriptional state of both cancer and immune cells in the tumor microenvironment that more accurately reflects metastatic propensity than does mutation analysis alone (**Fig. 5**).

While the CCF_BAP1_ was of less prognostic value than anticipated, the 15-GEP SVM discriminant score provided unexpected new insights into early tumor evolution and prognosis. We found that low discriminant score on either the Class 1 or Class 2 side of the decision boundary may indicate tumors in transition between these two states (**Fig. 5**). As such, a low discriminant score does not necessarily indicate “low confidence” but rather, it functions as a prognostic modifier associated with worse prognosis in Class 1 tumors and better prognosis in Class 2 tumors (**Fig. 4d-e**). A limitation of the chromosome copy number calling method is that it did not allow for precise CCF determination, such that LOH3p was assumed to be at ∼100% CCF. However, this assumption is reasonable based on previous findings^14,20,22^. Further work is warranted to determine how best to incorporate the discriminant score into precision clinical management, perhaps by identifying small tumors in transition that should be treated promptly rather than observed.

It has been suggested that *GNA11* is a “more potent” oncogene than *GNAQ* mutations because *GNA11* mutations may be detected more frequently in metastatic tumors ^10,36^. However, the present study does not support this claim. Whereas *GNA11* mutations were associated with some high-risk features such as increased patient age and increased tumor size, they were not associated with MFS and only weakly associated with OS. Further, *GNA11* mutations were associated with *BAP1* mutation status and Class 2 status, but they were rendered non-significant for both MFS and OS when either *BAP1* mutation status or 15-GEP were entered into a multivariate Cox model. Thus, *GNA11* mutations are associated with other high-risk features but do not appear to have independent prognostic significance and do not appear to be more potent than GNAQ mutations. Of further interest regarding G_q_ mutations, these were mutually exclusive as expected in all except 6 cases, which were of particular interest. In these 6 cases, a canonical G_q_ hotspot mutation – *GNAQ*^Q209P^, *GNA11*^Q209L^, *GNAQ*^R183Q^ or *GNA11*^R183C^ – was accompanied by a rare G_q_ pathway mutation – *GNAQ*^P193T^, *GNAQ*^T175M^, *CYSLTR2*^S154N^ or *PLCB4*^D630N^ (**Supplementary Table 2**). In 4 of these cases, the rare mutation was present at a higher variant allele frequency (VAF) than the hotspot mutation, suggesting that they occurred first but may have left residual selective pressure that led to the acquisition of another oncogenic G_q_ mutation. If *GNA11* mutations were more potent than *GNAQ* mutations, we hypothesize that cases might be found in which a *GNAQ* mutation was followed by a *GNA11* mutation, but none were detected. While we did not find prognostic value for G_q_ mutations independent of 15-GEP/PRAME, we did demonstrate the value of using the G_q_ mutation VAF to estimate tumor purity (**Supplementary Fig. 7a-b**). A potential limitation of this method is the inability to detect whole genome doubling, which could potentially skew the VAF of heterozygous mutations. However, whole genome doubling is rare in uveal melanoma and limited to a small minority (<7%) of large, advanced cases ^22^. Since our study comprised less than 10% of such advanced cases, this limitation is unlikely to have influenced our findings or conclusions.

In summary, this study confirms the prognostic value of UMAMs but demonstrates the inferiority of mutational analysis to the 15-GEP/PRAME classifier for prognostication. Nevertheless, UMAMs are relatively uncommon in other cancer types and can be useful for confirming the diagnosis of UM, which can be difficult in centers without specialized ocular cytopathology expertise ^37^. The most unexpected finding was the value of the SVM discriminant score for inferring the evolutionary state of small tumors in transition between Class 1 and Class 2, which moves us closer to a quantitative molecular method for inferring the malignant potential of uveal melanocytic tumors that straddle the line between benign nevus and small melanoma – a subject of considerable controversy in the field^30,38,39^. These findings are timely in light of prevailing evidence suggesting that UMs may metastasize when they are small and difficult to distinguish from benign nevi^24,30,40,41^, which could explain the failure of primary tumor treatment to prevent metastasis. Based on these results, a new prospective study is being planned to determine whether the discriminant score can be used in conjunction with the 15-GEP/PRAME classifier to guide the precision management of small uveal melanocytic tumors of indeterminate malignant potential by identifying lesions that are of sufficient risk of micrometastasis to warrant prompt treatment while sparing the vastly more abundant benign nevi that overlap in size^42^. Further studies and longer follow-up of this cohort will be important to further refine these prognostic tools for precision patient management.

## METHODS

### Patient Enrollment

Between January 2017 and April 2020, COOG2 enrolled 1687 subjects with UM involving the choroid, ciliary body and/or iris across 26 ocular oncology centers in the U.S. and Canada and prospectively monitored these subjects for metastatic progression and outcome. Informed consent was obtained from each patient. Primary treatment was performed according to the standard at each center. Federal Wide Assurance (FWA) from the Office of Human Research Protections (OHRP) and Institutional Review Board (IRB) or Ethics Committee approval was obtained in accordance with policies at each center. Exclusion criteria included patient age less than 18 years, diagnosis of a uveal tumor other than UM (e.g., metastatic cancer), prior radiotherapy, inadequate sample for molecular analysis, and patient withdrawal from the study. Prior photodynamic therapy or transpupillary thermotherapy were allowed if there was evidence of tumor regrowth. No participants were excluded based on sex, ethnicity, or race. Gender was recorded from medical records and used as a proxy for biological sex in this study. No additional data on gender identity was collected. For this analysis, a data lock was performed on March 4, 2024. Subjects were not included for this report if they had a primary iris melanoma (n=101), lacked adequate residual biopsy material for successful sequencing (n=212) or had no detectable UMAM (n=234).

### Tumor Sample Analysis

Prior to primary treatment, all subjects underwent clinical prognostic testing of their primary tumor using the DecisionDx^®^-UM 15-gene expression profile (15-GEP), which renders a result of Class 1 or Class 2, and DecisionDx^®^-PRAME, which renders a result of negative or positive. The 15-GEP test employs support vector machine (SVM) to classify each sample as Class 1 or Class 2 and to assign a discriminant score as a measure of confidence in the Class call based on the distance of a given sample to the decision boundary^32^. The residual material (∼25% of each clinical sample) was reserved for analysis on the custom UMAM NGS panel DecisionDx®-UMSeq, which has been described elsewhere^28^. This testing was performed in a CAP-accredited, CLIA-certified clinical laboratory (Castle Biosciences, Inc., Friendswood, TX, USA), as previously described^4,43^

Variants were sequenced with Ion GeneStudio S5 Prime Sequencer (ThermoFisher Scientific, Waltham, MA, USA) and processed with Ion Reporter (Version 5.6) software. Variant detection, analysis, and annotation was conducted with Ion Torrent Suite Browser (Version 5.8) and Ion Reporter using human reference sequence hg19. Sequencing quality assessment was conducted for each run, including total yield, useable reads, percent polyclonal reads, and amplicon coverage, as previously described^28^. Sample-specific sequencing quality metrics are included with **Supplementary Table 1**.

Mutations were classified as nonsense (introduction of a premature stop codon), stop-loss or start-loss (loss of stop or start codon preventing translation), frameshift insertion or deletion (shift of codon reading frame via addition or subtraction of a non-triplet set of nucleotides), non-frameshift insertion or deletion (addition or removal of a codon without shifting the reading frame), block substitution (alteration of multiple sequential codons), splice site alteration (alteration of splice donor or acceptor site), and missense (substitution of one amino acid). All of the following variants were called pathogenic: nonsense, stop-loss, start-loss, frameshift and non-frameshift insertions and deletions, and block substitutions. Splice site alterations were called pathogenic if predicted to result in splice acceptor or donor site loss or gain variant as predicted by a SpliceAI score greater than or equal to 0.5 ^44^. Missense variants were called pathogenic if they: (1) were previously reported as pathogenic in the ClinVar Database^45^, (2) exhibited a SIFT score less than or equal to 0.05, or (3) exhibited a PolyPhen2 score greater than or equal to 0.5. All genetic variants that were called pathogenic were classified as tier I, II, or III according to the guidelines of the College of American Pathologists (CAP), American Society of Clinical Oncology (ASCO), and Association for Molecular Pathology (AMP)^46^.

### Functional Assessment of *BAP1* Mutations Using Saturation Genome Editing Database

*BAP1* mutations involving complex alterations (≥5 nucleotide changes) were excluded from analysis and the remainder were converted from hg19 to hg38 reference genomes using the Broad Institute *Liftover* tool (https://liftover.broadinstitute.org/). Mutations were mapped to a CRISPR-based SGE database for BAP1, matching mutations based on hg38 start position, reference allele(s), and mutant allele(s) to retrieve the corresponding SGE functional classifications and scores as previously described^33^. Significance of functional classification was determined by two-tailed Fisher’s exact test, and significance of functional scores was determined by two-tailed Wilcoxon signed-rank test.

### Calculation of Tumor Purity, Variant Allele Frequency, and Cancer Cell Fraction

Tumor purity (TP), the percentage of cells in a sample that are tumor cells, was inferred from the VAF of the G_q_ mutation, assuming that the G_q_ mutation is the founder mutation, is a heterozygous alteration, and is therefore present at 50% VAF in tumor cells. In rare cases with more than one G_q_ mutation, the mutation with the highest frequency (and presumably the earlier initiating mutation) was used. As such, TP=*min*([VAF_Gq-mutant_ x 2],100%). To validate the estimation of tumor purity based on VAF of G_q_ mutations, we compared tumor purity estimation using VAF of G_q_ mutation to that using chromosome copy number variations in the UM TCGA cohort ^22^ using ABSOLUTE and FACETS. Statistical significance was determined using Pearson correlation (**Supplementary Fig. 7**).

The VAF for *BAP1*, *SF3B1*, and *EIF1AX* mutations was corrected for TP using the following equation: TP-corrected VAF_BSE=_VAF_BSE_ ÷ TP. Samples without a detectable G_q_ mutation could not be corrected for VAF and, thus, were not included in analyses requiring TP-corrected VAF_BSE_. Next, we estimated the cancer cell fraction (CCF) for each BSE mutation, representing the proportion of UM cells that harbor a given mutation, which requires a correction for allelic copy number. *SF3B1* is located on chromosome 2, which is not frequently altered in UM^20,22^. Thus, *SF3B1* mutations were assumed to be heterozygous and CCF*_SF3B1_*=*min*(TP-corrected VAF_SF3B1_ x 2, 100%). *EIF1AX* is located on the X chromosome, which is also rarely lost in UM^22^. Thus, gender was used to calculate mutant CCF_EIF1AX_, where females were assumed to have an *EIF1AX* mutation at 50% and males at 100% of TP-corrected VAF. Thus, the CCF*_EIF1AX_* for females was calculated as CCF*_EIF1AX_*=*min*(TP-corrected VAF_EIF1AX_ x 2, 100%), whereas the CCF*_EIF1AX_* for males was assumed to be equal to TP-corrected VAF_EIF1AX_. *BAP1* is located at chromosome 3p21^47^, which frequently undergoes copy number loss in UM ^20,22^. Thus, to detect loss of heterozygosity (LOH) and calculate CCF for *BAP1*, we developed a custom targeted CNV sequencing panel containing 74 loci across chromosome 3p that was performed on the same sample used for the 15-GEP/PRAME classifier and UMAM NGS panel. For *BAP1*-mutant tumors with retention of heterozygosity for chromosome 3p, the CCF*_BAP1_* was calculated as CCF*_BAP1_*=*min*(TP-corrected VAF_BAP1_ x 2, 100%). For tumors demonstrating LOH for chromosome 3p (LOH3p), CCF*_BAP1_* was assumed to be equal to TP-corrected VAF_BAP1_.

For the custom CNV sequencing panel, B-allele frequencies and log fold-change (lfc) read depths across chromosome 3p were compared to a reference DNA panel of normals (PON), comprising peripheral blood mononuclear cell (PBMC) samples from 64 patients. Variant call format (VCF) files were analyzed using Wheeljack (https://github.com/covingto/KRCGTK/releases/tag/v0.1). Copy-number loss for chromosome 3p was detected by consistent b-allele frequencies at 100% and a decreased lfc read depth of less than 0. Isodisomy for chromosome 3p was identified by consistent b-allele frequencies at 100% and a lfc read depth of approximately 0. For downstream analyses, samples demonstrating either copy number loss or isodisomy for chromosome 3p were called as LOH3p, whereas samples without these aberrations were called as retention of heterozygosity for 3p. Calls were made by hand and adjudicated by 3 of the authors (J.J.D, C.L.D., K.R.C.). Variability across b-allele and read depth plots was used to assign confidence scores with 0, 1, 2, and 3 corresponding to very low, low, medium, and high confidence, respectively. A confidence score of 2 or 3 was required for use in downstream analyses.

### Data Management

REDCap (https://projectredcap.org/), a secure HIPAA compliant application^48^, was used for electronic data management, as previously described^6^. Baseline data included date of enrollment, date and method of biopsy, cytology result (if available), date and method of primary tumor treatment, patient age at study entry, sex, self-reported race and ethnicity, iris color (blue/green, intermediate, or brown), tumor diameter, tumor thickness, ciliary body involvement, and metastatic status. The American Joint Committee on Cancer (AJCC) 8^th^ edition^49^ was used for tumor staging. Follow-up data included local tumor recurrence (tumor regrowth in the eye or orbit following radiotherapy or in the orbit following enucleation), metastatic status, date and location of initial metastasis, systemic status at last follow-up, and date and cause of death. Molecular test results were entered into REDCap by Castle Biosciences, which was masked to other REDCap data. Each center was masked to data entered by other centers and by Castle Biosciences. Only the coordinating center and COOG2 Data Committee had access to all data.

Baseline and follow-up ophthalmic visits were performed as per standard of care at each center but typically included a comprehensive ophthalmic examination, fundus photography, optical coherence tomography, and ultrasonography performed at least every 3-4 months for the first year after treatment, every 4-6 months for the second year, and every 6-12 months thereafter. Baseline systemic imaging was typically performed with CT of the chest, abdomen, and pelvis. Subsequent systemic surveillance typically included imaging of the liver with CT, MRI or ultrasound at least twice a year, along with chest CT or chest x-ray at least once a year.

### Statistical Analysis

Statistical analysis was performed using SAS 9.4 (SAS Institute, Cary, NC) and R (v4.2.2). Chi-square test was used to compare categorical variables unless expected frequencies were less than 5 for at least 25% of category cells, in which case Fisher exact test was used. Two-tailed Wilcoxon signed-rank test was used for comparing continuous variables. Statistical analysis of patient demographics and tumor characteristics for association with UMAMs compared all patients with a given mutation to all those without the mutation. All statistical tests were two-sided, and statistical significance was defined as *P<*0.05. Differences in metastasis-free survival (MFS, time from primary tumor treatment to initial detection of metastatic disease) and overall survival (OS, time from primary tumor treatment to death from any cause) associated with a given factor were evaluated using Kaplan-Meier (KM) survival curves and the log-rank test.

Propensity scores were calculated based on tumor thickness and tumor diameter to compare MFS and OS in Class 1 tumors that were wildtype versus mutant for *BAP1*, using a 3:1 matching ratio. Cox regression was used to assess the contribution of multiple factors influencing metastatic risk. Univariable and multivariable Cox models were constructed to assess the impact of variables both separately and in combination. Survival analysis for continuous variables (e.g., discriminant score) was performed by calculating survival probabilities at specified time points using a time-to-event model that includes the continuous variable^50^.

## DATA AVAILABILITY

Relevant data available in Supplementary Information. Raw sequencing data are accessible via database of Genotypes and Phenotypes (dbGaP) under accession number TBA-XXXXX. Access to the data requires an approved application through dbGaP due to patient privacy concerns. In addition to the data available within the Supplementary Information, data is publicly available at the Dryad Research Data Repository: https://doi.org/10.5061/dryad.z8w9ghxqk. Other relevant data supporting the findings of this study are available from the corresponding author upon request.

## CODE AVAILABILITY

Custom code used for calculations, including tumor purity and cancer cell fraction, as well as for visualization, is available at https://github.com/jwharbour/COOG2tools. The repository includes scripts for preprocessing, statistical analysis, and generating figures. For inquiries or assistance, please contact the corresponding author.

## ACKNOWLEDGMENTS

This research was supported by National Eye Institute to J.W.H. (R01 CA125970); Cancer Prevention and Research Institute of Texas Recruitment of Established Investigator Award to J.W.H. (RR220010); Research to Prevent Blindness, Inc. Senior Scientific Investigator Award to J.W.H.; National Cancer Institute Support Grants to Simmons Comprehensive Cancer Center, University of Texas Southwestern Medical Center (P30 CA142543) and Sylvester Comprehensive Cancer Center, University of Miami (P30 CA240139); National Eye Institute Core Grants to the Department of Ophthalmology, University of Texas Southwestern Medical Center (P30 EY030413), Bascom Palmer Eye Institute and Department of Ophthalmology, University of Miami Department of Ophthalmology (P30 EY014801) and Casey Eye Institute, Oregon Health & Science University (P30 EY010572); Research to Prevent Blindness, Inc. Challenge Grant to the Department of Ophthalmology, University of Texas Southwestern Medical Center; Research to Prevent Blindness, Inc. Unrestricted Grants to Bascom Palmer Eye Institute, Department of Ophthalmology, University of Miami and Casey Eye Institute, Oregon Health & Science University; Malcolm M. Marquis, MD Endowed Fund for Innovation to Casey Eye Institute, Oregon Health & Science University; and Castle Biosciences, Inc. grant to University of Miami. The authors thank our patients who generously participated in this project. We acknowledge Katherina M. Alsina, PhD, Jason H. Rogers, MS, Jennifer J. Siegel, PhD, Jeff Wilkinson, PhD, Lauren Sholl, MS, and Daniel Vargas, MS, from Castle Biosciences, Inc., for data management support, and the following individuals for their dedication to accurate data entry at each study site: Angie Adler, Mutaz Al-Nawaflh, Corrina Azarcon, Buse Guneri Beser, Karina Bostwick, Nury Cabrera, Teja Chemudupati, Caroline Craven, Jessica Fitch, Nancy Gee, Ashley Go, Caleb Hartley, Mustafa Hashmi, Tyler Hendrickson, Gary Lamoureux, Anne Marie Lane, Kiley Lazarek, Ashton Leone, Ronan McCarthy, Audra Miller, Trece Mayhan, Monica Oxenreiter, Barbara Perez, Dayana Pineda, Nicki Plocharsky, Mary Preston, Kourtney Storey, Laurie Tavernier, Bonnie Verges, Holly Vincent, Brooke Waller, and Aaron Yeung.

## Author contributions

J.W.H conceived of, designed, and acquired the financial support for the study. J.W.H., C.L.D., and Z.M.C. provided administrative support. E.W., A.C.S., M.A.M., T.S.F., A.H.S., D.A.R., I.K., K.D.P., H.D., T.A.A, P.M., B.K.W., E.S., S.C.N.O, D.H.C., A.C., J.M., S.D.W., M.A., J.W., D.S.G., J.D., P.H., T.T., C.J., Z.M.C., and J.W.H. provided study materials and enrolled patients for the study. J.J.D., C.L.D., and P.M. collected and assembled data. J.J.D., K.R.C., M.A.D., S.Z., E.W., and J.W.H. conducted data analysis and interpretation. J.J.D. and J.W.H. wrote the original draft of the manuscript. All authors provided critical input and contributed to discussions and revisions of the manuscript.

## REFERENCES

1 Carvajal, R. D. et al. Advances in the clinical management of uveal melanoma. Nat Rev Clin Oncol 20, 99–115 (2023). 10.1038/s41571-022-00714-1

2 Onken, M. D., Worley, L. A., Ehlers, J. P. & Harbour, J. W. Gene expression profiling in uveal melanoma reveals two molecular classes and predicts metastatic death. Cancer Res 64, 7205–7209 (2004).

3 Field, M. G. et al. PRAME as an independent biomarker for metastasis in uveal melanoma. Clin Cancer Res 22, 1234–1242 (2016). 10.1158/1078-0432.ccr-15-2071

4 Field, M. G. et al. Epigenetic reprogramming and aberrant expression of PRAME are associated with increased metastatic risk in Class 1 and Class 2 uveal melanomas. Oncotarget 7, 59209–59219 (2016). 10.18632/oncotarget.10962

5 Cai, L. et al. Gene Expression Profiling and PRAME Status Versus Tumor-Node-Metastasis Staging for Prognostication in Uveal Melanoma. Am J Ophthalmol 195, 154–160 (2018). 10.1016/j.ajo.2018.07.045

6 Harbour, J. W. et al. 15-Gene Expression Profile and PRAME as Integrated Prognostic Test for Uveal Melanoma: First Report of Collaborative Ocular Oncology Group Study No. 2 (COOG2.1). J Clin Oncol 42, 3319–3329 (2024). 10.1200/JCO.24.00447

7 Decatur, C. L. et al. Driver Mutations in Uveal Melanoma: Associations With Gene Expression Profile and Patient Outcomes. JAMA Ophthalmol 134, 728–733 (2016). 10.1001/jamaophthalmol.2016.0903

8 Onken, M. D. et al. Oncogenic mutations in GNAQ occur early in uveal melanoma. Investigative ophthalmology & visual science 49, 5230–5234 (2008). 10.1167/iovs.08-2145

9 Van Raamsdonk, C. D. et al. Frequent somatic mutations of GNAQ in uveal melanoma and blue naevi. Nature 457, 599–602 (2009). nature07586 [pii] 10.1038/nature07586 [doi]

10 Van Raamsdonk, C. D. et al. Mutations in GNA11 in uveal melanoma. N Engl J Med 363, 2191–2199 (2010). 10.1056/NEJMoa1000584

11 Johansson, P. et al. Deep sequencing of uveal melanoma identifies a recurrent mutation in PLCB4. Oncotarget 7, 4624–4631 (2016). 10.18632/oncotarget.6614

12 Moore, A. R. et al. Recurrent activating mutations of G-protein-coupled receptor CYSLTR2 in uveal melanoma. Nature genetics 48, 675–680 (2016). 10.1038/ng.3549

13 Vader, M. J. C. et al. GNAQ and GNA11 mutations and downstream YAP activation in choroidal nevi. Br J Cancer 117, 884–887 (2017). 10.1038/bjc.2017.259

14 Durante, M. A. et al. Genomic evolution of uveal melanoma arising in ocular melanocytosis. Cold Spring Harb Mol Case Stud 5 (2019). 10.1101/mcs.a004051

15 Solomon, D. A. et al. Iris and Ciliary Body Melanocytomas Are Defined by Solitary GNAQ Mutation Without Additional Oncogenic Alterations. Ophthalmology 129, 1429–1439 (2022). 10.1016/j.ophtha.2022.07.002

16 Harbour, J. W. et al. Frequent mutation of BAP1 in metastasizing uveal melanomas. Science 330, 1410–1413 (2010). 10.1126/science.1194472

17 Harbour, J. W. et al. Recurrent mutations at codon 625 of the splicing factor SF3B1 in uveal melanoma. Nature genetics 45, 133–135 (2013). 10.1038/ng.2523

18 Furney, S. J. et al. SF3B1 mutations are associated with alternative splicing in uveal melanoma. Cancer Discov 3, 1122–1129 (2013). 10.1158/2159-8290.CD-13-0330

19 Martin, M. et al. Exome sequencing identifies recurrent somatic mutations in EIF1AX and SF3B1 in uveal melanoma with disomy 3. Nature genetics 45, 933–936 (2013). 10.1038/ng.2674

20 Field, M. G. et al. Punctuated evolution of canonical genomic aberrations in uveal melanoma. Nat Commun 9, 116 (2018). 10.1038/s41467-017-02428-w

21 Carbone, M. et al. Biological Mechanisms and Clinical Significance of BAP1 Mutations in Human Cancer. Cancer Discov 10, 1103–1120 (2020). 10.1158/2159-8290.CD-19-1220

22 Robertson, A. G. et al. Integrative analysis identifies four molecular and clinical subsets in uveal melanoma. Cancer Cell 32, 204–220 e215 (2017). 10.1016/j.ccell.2017.07.003

23 Durante, M. A. et al. Single-cell analysis reveals new evolutionary complexity in uveal melanoma. Nat Commun 11, 496 (2020). 10.1038/s41467-019-14256-1

24 Eskelin, S., Pyrhonen, S., Summanen, P., Hahka-Kemppinen, M. & Kivela, T. Tumor doubling times in metastatic malignant melanoma of the uvea: tumor progression before and after treatment. Ophthalmology 107, 1443–1449 (2000). 10.1016/s0161-6420(00)00182-2

25 Aronow, M. E., Topham, A. K. & Singh, A. D. Uveal Melanoma: 5-Year Update on Incidence, Treatment, and Survival (SEER 1973-2013). Ocular oncology and pathology 4, 145-151 (2018). 10.1159/000480640

26 Rodriguez, D. A. et al. Multiregional genetic evolution of metastatic uveal melanoma. NPJ Genom Med 6, 70 (2021). 10.1038/s41525-021-00233-5

27 Shain, A. H. et al. The genetic evolution of metastatic uveal melanoma. Nature genetics 51, 1123–1130 (2019). 10.1038/s41588-019-0440-9

28 Alsina, K. M. et al. Analytical Validation and Performance of a 7-Gene Next-Generation Sequencing Panel in Uveal Melanoma. Ocular oncology and pathology 7, 428–436 (2021). 10.1159/000518829

29. Harbour, J. W. & Shih, H. A. in UpToDate, Waltham, MA. (Accessed on April 19, 2022). (eds M.B. Atkins & R.S. Berman) (2018).

30 Harbour, J. W. et al. Are Risk Factors for Growth of Choroidal Nevi Associated With Malignant Transformation? Assessment With a Validated Genomic Biomarker. Am J Ophthalmol 197, 168–179 (2019). 10.1016/j.ajo.2018.08.045

31 Walter, S. D. et al. Prognostic Implications of Tumor Diameter in Association With Gene Expression Profile for Uveal Melanoma. JAMA Ophthalmol 134, 734–740 (2016). 10.1001/jamaophthalmol.2016.0913

32 Plasseraud, K. M. et al. Gene expression profiling in uveal melanoma: technical reliability and correlation of molecular class with pathologic characteristics. Diagn Pathol 12, 59 (2017). 10.1186/s13000-017-0650-3

33 Waters, A. J. et al. Saturation genome editing of BAP1 functionally classifies somatic and germline variants. Nature genetics 56, 1434–1445 (2024). 10.1038/s41588-024-01799-3

34 Iyer, S. V., Goodwin, S. & McCombie, W. R. Leveraging the power of long reads for targeted sequencing. Genome research 34, 1701–1718 (2024). 10.1101/gr.279168.124

35 Kaler, C. J. et al. BAP1 Loss Promotes Suppressive Tumor Immune Microenvironment via Upregulation of PROS1 in Class 2 Uveal Melanomas. Cancers (Basel) 14 (2022). 10.3390/cancers14153678

36 Terai, M. et al. Prognostic Values of G-Protein Mutations in Metastatic Uveal Melanoma. Cancers (Basel) 13 (2021). 10.3390/cancers13225749

37 Correa, Z. M. & Augsburger, J. J. Sufficiency of FNAB aspirates of posterior uveal melanoma for cytologic versus GEP classification in 159 patients, and relative prognostic significance of these classifications. Graefes Arch Clin Exp Ophthalmol 252, 131–135 (2014). 10.1007/s00417-013-2515-0

38 Harbour, J. W., Correa, Z. M. & Stacey, A. W. Do Short Delays in Treatment Affect Uveal Melanoma Prognosis? Ophthalmology (2024). 10.1016/j.ophtha.2024.05.018

39 Singh, A. D., Raval, V., Wrenn, J. & Zabor, E. C. Small Choroidal Melanoma: Outcomes After Surveillance Versus Immediate Treatment. Am J Ophthalmol 241, 47–56 (2022). 10.1016/j.ajo.2022.03.024

40 Nichols, E. E., Richmond, A. & Daniels, A. B. Micrometastatic Dormancy in Uveal Melanoma: A Comprehensive Review of the Evidence, Mechanisms, and Implications for Future Adjuvant Therapies. Int Ophthalmol Clin 57, 1–10 (2017). 10.1097/iio.0000000000000160

41 Augsburger, J. J. et al. Diagnostic transvitreal fine-needle aspiration biopsy of small melanocytic choroidal tumors in nevus versus melanoma category. Trans Am Ophthalmol Soc 100, 225–232; discussion 232-224 (2002).

42 Augsburger, J. J., Correa, Z. M., Trichopoulos, N. & Shaikh, A. Size overlap between benign melanocytic choroidal nevi and choroidal malignant melanomas. Investigative ophthalmology & visual science 49, 2823–2828 (2008). 10.1167/iovs.07-1603

43 Onken, M. D., Worley, L. A., Tuscan, M. D. & Harbour, J. W. An accurate, clinically feasible multi-gene expression assay for predicting metastasis in uveal melanoma. J Mol Diagn 12, 461–468 (2010). 10.2353/jmoldx.2010.090220

44 Jaganathan, K. et al. Predicting Splicing from Primary Sequence with Deep Learning. Cell 176, 535–548 e524 (2019). 10.1016/j.cell.2018.12.015

45 Landrum, M. J. et al. ClinVar: public archive of relationships among sequence variation and human phenotype. Nucleic acids research 42, D980–D985 (2014). 10.1093/nar/gkt1113

46 Li, M. M. et al. Standards and Guidelines for the Interpretation and Reporting of Sequence Variants in Cancer: A Joint Consensus Recommendation of the Association for Molecular Pathology, American Society of Clinical Oncology, and College of American Pathologists. J Mol Diagn 19, 4–23 (2017). 10.1016/j.jmoldx.2016.10.002

47 Jensen, D. E. et al. BAP1: a novel ubiquitin hydrolase which binds to the BRCA1 RING finger and enhances BRCA1-mediated cell growth suppression. Oncogene 16, 1097–1112 (1998).

48 Harris, P. A. et al. Research electronic data capture (REDCap)--a metadata-driven methodology and workflow process for providing translational research informatics support. Journal of biomedical informatics 42, 377–381 (2009). 10.1016/j.jbi.2008.08.010

49. AJCC Ophthalmic Oncology Task Force. International Validation of the American Joint Committee on Cancer’s 7th Edition Classification of Uveal Melanoma. JAMA Ophthalmol 133, 376-383 (2015). 10.1001/jamaophthalmol.2014.5395

50 Denz, R. & Timmesfeld, N. Visualizing the (Causal) Effect of a Continuous Variable on a Time-To-Event Outcome. Epidemiology 34, 652–660 (2023). 10.1097/EDE.0000000000001630

