## Supplementary Information for "Early Genetic Evolution of Driver Mutations in Uveal Melanoma"

**Supplementary Table 1.** Summary of clinical and demographic features (n=1140).

| Features | Values |
|---|---|
| Age at study entry, years | |
| Median (range) | 64.3 (18-99) |
| Mean (SD) | 62.5 (13.63) |
| Male sex, No. (%) | 590 (51.75) |
| Ethnicity, No. (%) | |
| Non-Hispanic or Latino | 1075 (94.3%) |
| Hispanic or Latino | 41 (3.6%) |
| Not specified | 24 (2.1%) |
| Race, No. (%) | |
| Caucasian | 1095 (96.3%) |
| Black | 8 (0.7%) |
| Asian | 6 (0.5%) |
| Native American/Alaskan | 2 (0.2%) |
| More than one race | 3 (0.3%) |
| Not specified | 26 (2.3%) |
| Eye, Right, No. (%) | 578 (50.7%) |
| Iris color, No. (%) | |
| Blue/green | 453 (39.7%) |
| Brown | 167 (14.7%) |
| Intermediate | 89 (7.8%) |
| Not specified | 431 (37.8%) |
| Ciliary body involvement, No. (%) | 201 (17.6%) |
| Tumor diameter, mm | |
| Median (range) | 12.1 (3-29) |
| Mean (SD) | 12.6 (+/-3.9) |
| Tumor thickness, mm | |
| Median (range) | 4.5 (1-18) |
| Mean (SD) | 5.5 (+/-3.1) |
| 15-GEP, No. (%) | |
| Class 1 | 716 (62.8%) |
| Class 2 | 424 (37.2%) |
| PRAME, No. (%) | |

| | |
|---|---|
| Negative (–) | 757 (66.4%) |
| Positive (+) | 383 (33.6%) |
| Death, No. (%) | 241 (21.1%) |
| Time to death or last follow-up, months | |
| Median (range) | 52.8 (0-101.7) |
| Mean (SD) | 52.4 (+/- 22.1) |
| Metastasis, No. (%) | 229 (20.1%) |
| Time to metastasis or last follow-up, months | |
| Median (range) | 50.8 (0-101.7) |
| Mean (SD) | 49.1 (+/- 24.0) |
| Local recurrence, No. (% of all patients) | 54 (4.7%) |
| Time to local recurrence, months | |
| Median (range) | 28.5 (3.5-82.2) |
| Mean (SD) | 30.2 (18.6) |
| Metastasis after local recurrence, No. (%) | 28 (51.9%) |

Abbreviations: 15-GEP, 15-gene expression profile test; SD, standard deviation; No., number.

**Supplementary Table 2. Cohort Summary.** Table of patient annotations, including clinical features and outcomes, uveal melanoma-associated mutations, and chromosome 3p status (n=1140). Table available in excel file.

**Supplementary Table 3. Clinical features associated with uveal melanoma-associated mutations.** Table providing the statistical analysis of uveal melanoma-associated mutations (n=1140). Continuous variables were analyzed by two-tailed Wilcoxon rank-sum test and discrete variables by chi-squared test or Fisher test if indicated by †.
Abbreviations: 15-GEP, 15-gene expression profile; *PRAME*(+/-), positive/negative *PRAME* expression; No., number; *GNAQ*, *GNA11*, *PLCB4*, *CYSLTR2*, *BAP1*, *SF3B1* and *EIF1AX* indicate pathogenic mutations in these genes. Table available in excel file.

**Supplementary Table 4.** Univariate Cox regression analyses of metastasis-free survival and overall survival.*

| Risk Factor | Metastasis-Free Survival | | Overall Survival | |
|---|---|---|---|---|
| | Univariate HR (95% CI) | *P* | Univariate HR (95% CI) | *P* |
| 15-GEP Class 2 | 11.0 (7.8,15.5) | <.0001 | 6.3 (4.7,8.5) | <.0001 |
| *PRAME*(+) | 3.3 (2.5,4.2) | <.0001 | 2.9 (2.2,3.7) | <.0001 |
| *GNAQ* | 0.8 (0.6,1.0) | 0.05 | 0.7 (0.5,0.9) | 0.001 |
| *GNA11* | 1.1 (0.9,1.5) | 0.4 | 1.4 (1.1,1.9) | 0.006 |
| *PLCB4* | 1.5 (0.7,3.3) | 0.3 | 1.2 (0.5,2.6) | 0.7 |
| *CYSLTR2* | 2.0 (0.8,4.9) | 0.1 | 0.7 (0.2,2.9) | 0.6 |
| *BAP1* | 5.9 (4.5,7.8) | <.0001 | 4.3 (3.3,5.6) | <.0001 |
| *SF3B1* | 0.5 (0.3,0.7) | 0.0009 | 0.5 (0.3,0.7) | 0.0005 |
| *EIF1AX* | 0.2 (0.1,0.3) | <.0001 | 0.4 (0.3,0.6) | <.0001 |
| $CCF_{BAP1}$* | 1.0 (1.0,1.0) | 1.0 | 1.0 (1.0,1.0) | 0.8 |
| $CCF_{SF3B1}$* | 1.03 (1.0,1.1) | 0.3 | 1.0 (1.0,1.1) | 0.8 |
| $CCF_{EIF1AX}$* | 1.00 (1.0,1.0) | 0.8 | 0.99 (1.0,1.0) | 0.1 |

*Includes the COOG2.2 cohort (n=1133), without cases that had metastasis at baseline (n=7). Abbreviations: CI, confidence interval; 15-GEP, 15-gene expression profile; PRAME(+), positive PRAME expression; HR, hazard ratio. *GNAQ*, *GNA11*, *PLCB4*, *CYSLTR2*, *BAP1*, *SF3B1* and *EIF1AX* indicate pathogenic mutations in these genes.

**Supplementary Table 5.** Multivariate Cox regression analysis of metastasis-free survival and overall survival.

| Risk Factor Pairs* | Metastasis-Free Survival | | Overall Survival | |
|---|---|---|---|---|
| | Multivariate HR (95% CI) | *P* | Multivariate HR (95% CI) | *P* |
| 15-GEP Class 2 | 10.1 (6.5,15.7) | <.0001 | 5.7 (3.8,8.6) | <.0001 |
| BAP1 | 1.1 (0.8,1.6) | 0.5 | 1.2 (0.8,1.7) | 0.5 |
| 15-GEP Class 2 | 12.8 (8.8,18.6) | <.0001 | 6.4 (4.7,8.8) | <.0001 |
| SF3B1 | 1.7 (1.1,2.8) | 0.03 | 1.1 (0.7,1.7) | 0.8 |
| 15-GEP Class 2 | 9.5 (6.6,13.8) | <.0001 | 6.5 (4.6,9.0) | <.0001 |
| EIF1AX | 0.6 (0.4,1.1) | 0.08 | 1.1 (0.7,1.6) | 0.8 |
| 15-GEP Class 2 | 11.2 (7.9,15.7) | <.0001 | 6.2 (4.6,8.3) | <.0001 |
| GNAQ | 1.1 (0.8,1.4) | 0.5 | 0.9 (0.7,1.1) | 0.3 |
| 15-GEP Class 2 | 11.2 (7.9,15.7) | <.0001 | 6.2 (4.6,8.3) | <.0001 |
| GNA11 | 0.9 (0.7,1.2) | 0.4 | 1.2 (0.9,1.5) | 0.2 |
| BAP1 | 5.9 (4.4,7.8) | <.0001 | 4.2 (3.2, 5.5) | <.0001 |
| GNAQ | 1.0 (0.7,1.2) | 0.7 | 0.8 (0.6,1.0) | 0.05 |
| BAP1 | 5.9 (4.5,7.8) | <.0001 | 4.2 (3.3, 5.5) | <.0001 |
| GNA11 | 1.0 (0.8,1.3) | 0.9 | 1.3 (1.0,1.7) | 0.05 |
| PRAME(+)** | 6.7 (3.2,14.2) | <.0001 | 2.7 (1.5,5.0) | 0.001 |
| SF3B1** | 0.8 (0.4,1.6) | 0.5 | 0.8 (0.4,1.4) | 0.4 |

*Includes the COOG2.2 cohort (n=1133), without cases that had metastasis at baseline (n=7).
**Class 1 only (n=715).
Abbreviations: CI, confidence interval; 15-GEP, gene expression profile; *PRAME*(+), positive *PRAME* expression; HR, hazard ratio; *GNAQ*, *GNA11*, *BAP1*, *SF3B1* and *EIF1AX* indicate pathogenic mutations in these genes. *PLCB4* and *CYSLTR2* not included due to small number of cases.

**Supplementary Table 6. Comparison of small versus larger tumors.** Table providing the statistical analysis of small tumors (n=131) versus larger tumors (n=1009). Continuous variables were analyzed by two-tailed Wilcoxon rank-sum test and discrete variables by chi-squared test or Fisher test if indicated (†). Table available in excel file.

**Supplementary Table 7. Comparison of BAP1 mutant versus BAP1 wild type uveal melanomas.** Continuous variables were analyzed by two-tailed Wilcoxon rank-sum test and discrete variables by chi-squared test or Fisher test if indicated by †. Table available in excel file.

**Supplementary Table 8. Comparison of clinical and molecular features associated with cancer cell fraction.** Table providing the statistical analysis of discrete variables associated with cancer cell fraction for patients with *BAP1* mutations and quality copy number calls (n=287), *SF3B1* (n=190), and *EIF1AX* mutations (n=295). Variables were analyzed by two-tailed Wilcoxon rank-sum test.

| Dichotomous Variable | | CCF | CCF | | |
|---|---|---|---|---|---|
| Group 1 | Group 2 | | Group 1* | Group 2* | *P*-value |
| Small Tumors | Larger Tumors | BAP1 | 87.8 ± 3.8 | 84.9 ± 1.1 | 0.3 |
| | | SF3B1 | 83.0 ± 4.4 | 94.3 ± 0.9 | 0.002 |
| | | EIF1AX | 89.9 ± 3.2 | 95.1 ± 0.8 | 0.04 |
| Class 1 | Class 2 | BAP1 | 79.6 ± 4.9 | 85.4 ± 1.1 | 0.2 |
| | | SF3B1 | 93.1 ± 0.9 | 96.5 ± 1.2 | 0.9 |
| | | EIF1AX | 94.4 ± 0.9 | 94.7 ± 2.1 | 0.2 |
| PRAME(-) | PRAME(+) | BAP1 | 85.5 ± 1.3 | 84.6 ± 1.8 | 0.9 |
| | | SF3B1 | 91.4 ± 2.0 | 94.1 ± 0.9 | 0.5 |
| | | EIF1AX | 94.4 ± 0.9 | 94.9 ± 1.5 | 0.09 |
| No CB Involvement | CB Involvement | BAP1 | 85.3 ± 1.2 | 84.4 ± 2.5 | 0.8 |
| | | SF3B1 | 92.5 ± 1.1 | 97.0 ± 0.6 | 0.8 |
| | | EIF1AX | 94.8 ± 0.8 | 90.5 ± 3.9 | 0.04 |

* Mean ± Standard Error

**Supplementary Table 9.** Univariate Cox regression analyses of metastasis-free survival and overall survival in Class 1 (n=467) and Class 2 (n=278) tumors without metastasis at baseline.

| **Mutated Gene** | **15-GEP Class** | **Metastasis-Free Survival** | | **Overall Survival** | |
|---|---|---|---|---|---|
| | | **Hazard Ratio (95% CI)** | ***P*** | **Hazard Ratio (95% CI)** | ***P*** |
| *BAP1** | Both Classes | 1.0 (1.0, 1.0) | 1.0 | 1.0 (1.0, 1.0) | 0.8 |
| | Class 1 | 1.0 (0.9, 1.0) | 0.5 | 1.0 (0.9,1.0) | 0.5 |
| | Class 2 | 1.0 (1.0, 1.0) | 0.8 | 1.0 (1.0,1.0) | 0.7 |
| *SF3B*** | Both Classes | 1.0 (1.0, 1.1) | 0.3 | 1.0 (1.0, 1.1) | 0.3 |
| | Class 1 | 1.0 (1.0, 1.1) | 0.3 | 1.0 (1.0, 1.1) | 0.3 |
| | Class 2 | 0.9 (0.6, 1.2) | 0.4 | 0.5 (0.04, 6.0) | 0.6 |
| *EIF1AX*** | Both Classes | 1.0 (1.0, 1.0) | 0.8 | 1.0 (1.0, 1.0) | 0.1 |
| | Class 1 | 1.0 (1.0, 1.0) | 1.0 | 1.0 (1.0, 1.0) | 0.1 |
| | Class 2 | 1.0 (0.9, 1.1) | 0.7 | 1.0 (1.0, 1.1) | 0.8 |

* Indicates samples with a *BAP1* mutation and evaluable chromosome 3 copy number call (confidence score 2 or 3) (n=283).

** Indicates samples with a respective *SF3B1* (n=189) or *EIF1AX* (n=295) mutation.

Abbreviations: MFS, metastasis-free survival; OS, overall survival; CI, confidence interval.

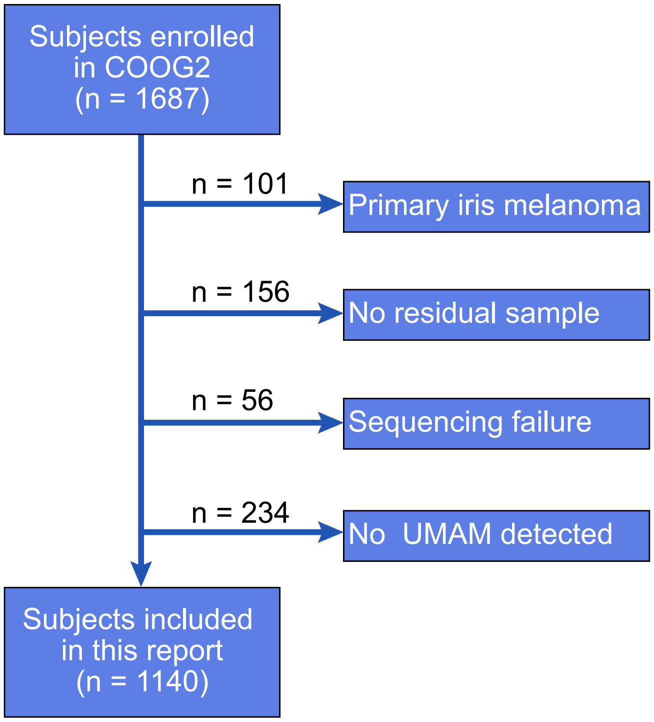


**Supplementary Figure 1.** Overview of subjects included in Collaborative Ocular Oncology Group Study 2 Report Number 2 (COOG2.2).

**a**

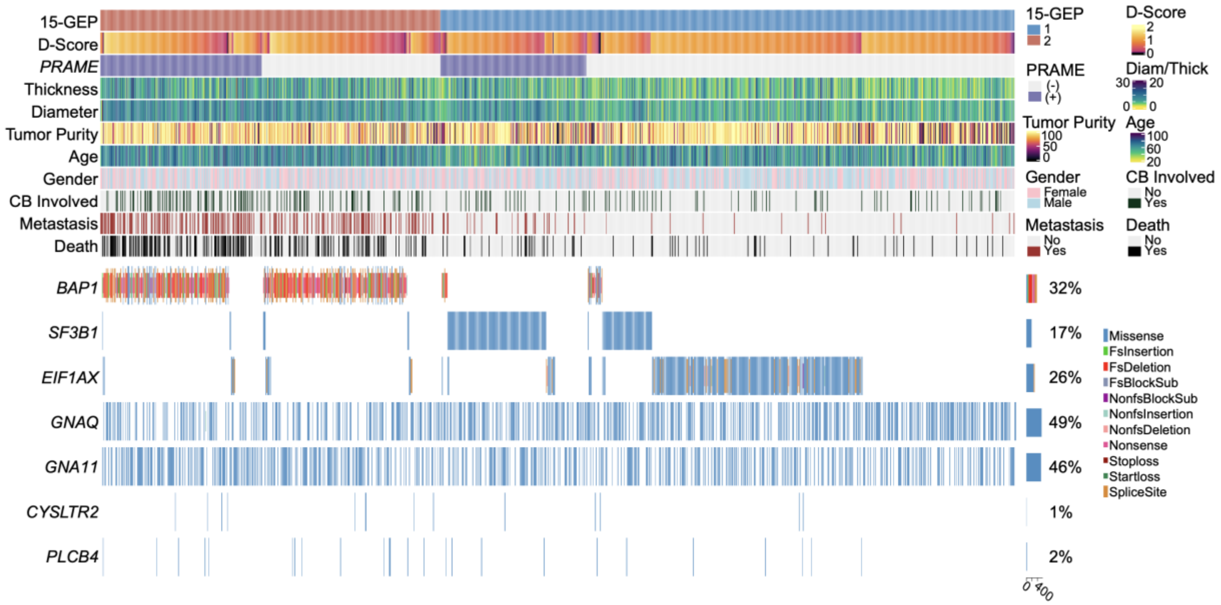


**Supplementary Figure 2.** Oncoprint for 1140 primary uveal melanomas with equivalent data as Figure 1, with samples sorted according to 15-GEP Class and PRAME status followed by *BAP1*, *SF3B1*, and *EIF1AX* mutation and decreasing discriminant score to emphasize the genetic landscape associated with 15-GEP Class and *PRAME* status. Diam, tumor diameter; Thick, tumor thickness. CB, ciliary body. D-score, 15-GEP support vector machine discriminant score. Variant types described in Materials and Methods.

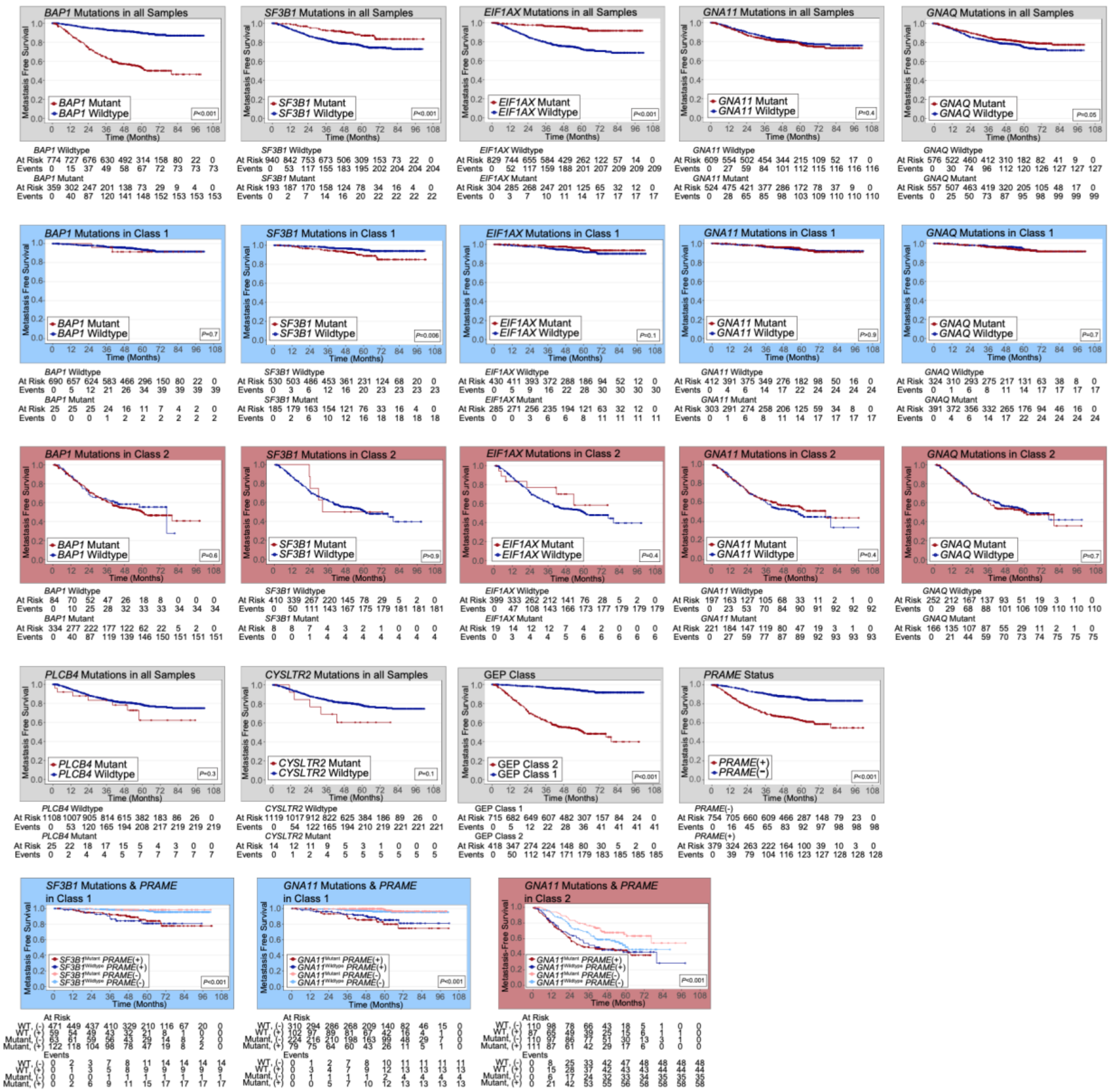


**Supplementary Figure 3.** Kaplan-Meier survival curves for metastasis-free survival in patients with detectable UMAMs in all samples (gray outline box) (n=1133), class 1 (blue outline box) (n=715), and class 2 (red outline box) (n=418). The last row depicts survival curves for *SF3B1* mutations or *GNA11* mutations stratified by *PRAME* status in Class 1 and Class 2 UM. Abbreviations: 15-GEP, 15-gene expression profile; MFS, metastasis-free survival.

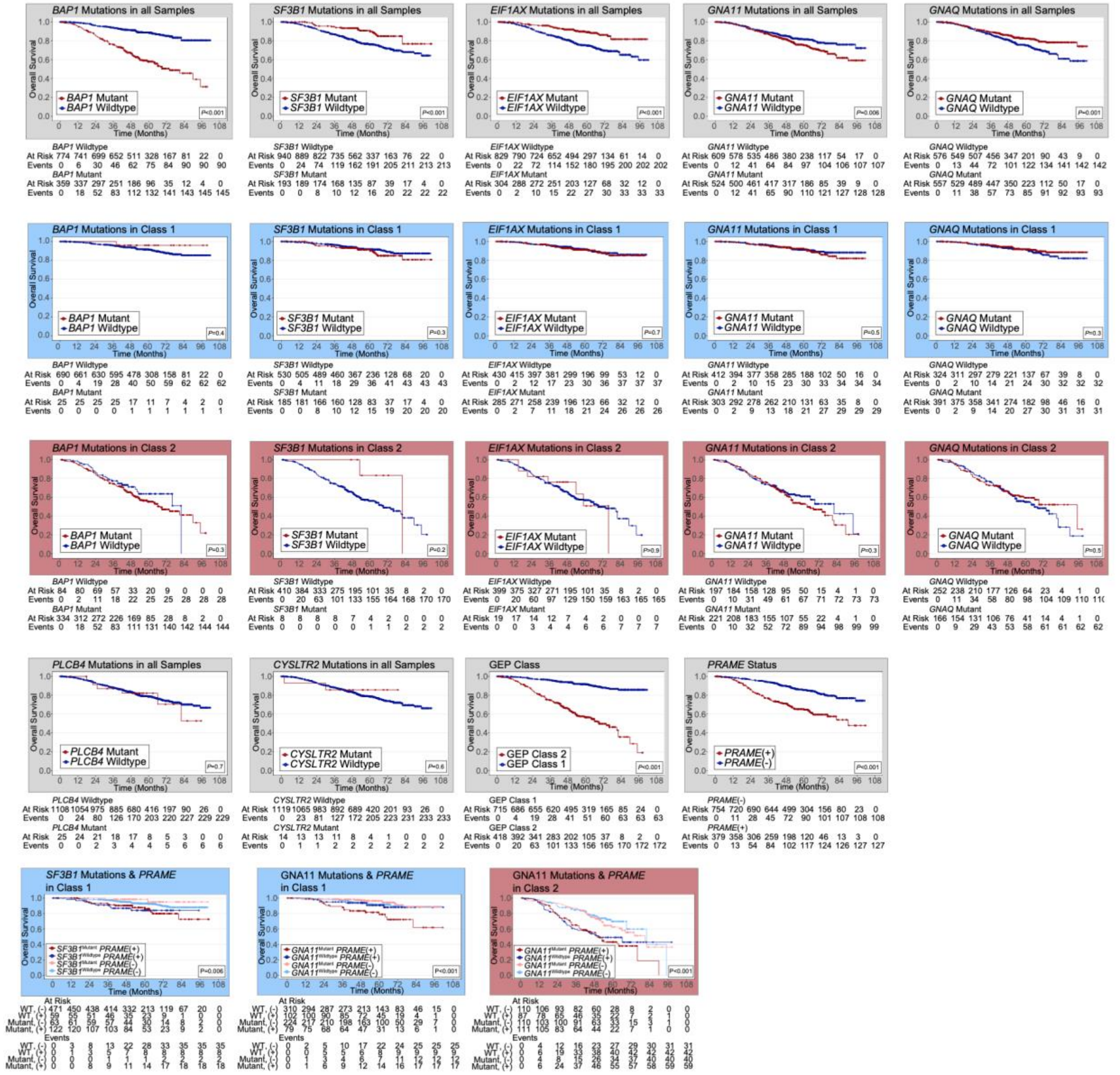


**Supplementary Figure 4.** Kaplan-Meier survival curves for overall survival in patients with detectable uveal melanoma-associated mutations in all samples (gray outline box) (n=1133), Class 1 (blue outline box) (n=715), and Class 2 (red outline box) (n=418). The last row depicts survival curves for *SF3B1* mutations or *GNA11* mutations stratified by *PRAME* status in Class 1 and Class 2 UM.

Abbreviations: OS, overall survival.

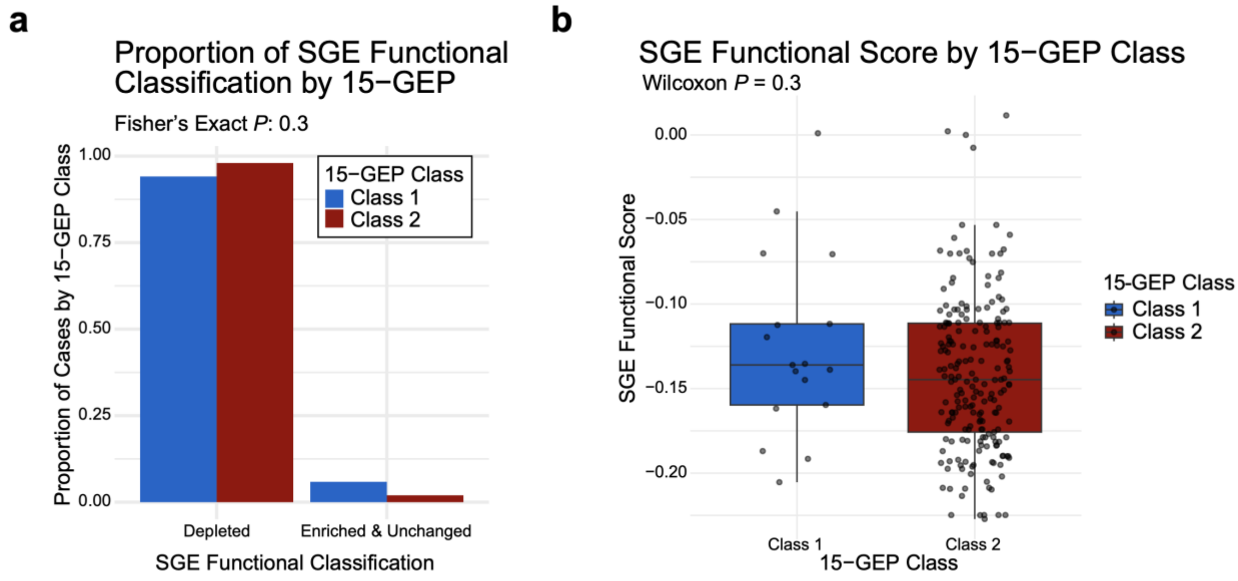


**Supplementary Figure 5.** Functional Assessment of *BAP1* Mutations (n=218) with CRISPR-based Saturation Genome Editing (SGE) Database published in Water et al., 2023, with comparison of 15-GEP Class 1 (n=17) versus Class 2 (n=201) tumors. **a**, Bar plot depicting the frequency of deleterious functional classification ("Depleted") compared to non-deleterious classification ("Enriched" or "Unchanged") for *BAP1* mutations according to 15-GEP Class status. **b**, Box plot exhibiting the SGE functional score of *BAP1* mutations for 15-GEP Class 1 and Class 2 tumors. Significance for comparing functional classifications and scores was determined by two-tailed Fisher's exact test and two-tailed Wilcoxon signed-rank test.

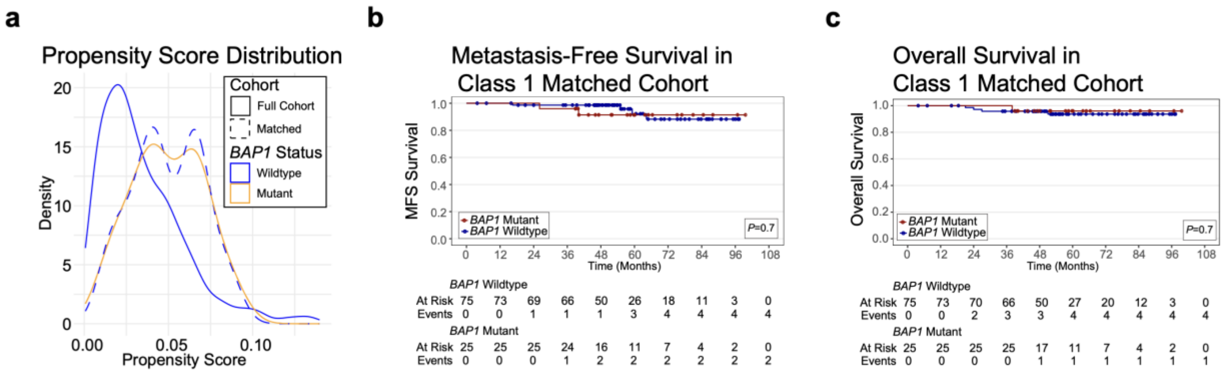


**Supplementary Figure 6.** Propensity score-matched survival analysis of Class 1 *BAP1* mutants (n=25) versus Class 1 *BAP1* wildtype (n=75) tumors, selected from the full cohort of Class 1 *BAP1* wildtype tumors without metastasis at baseline (n=690). **a**, Distribution of propensity scores for Class 1 tumors by *BAP1* mutation status and matching status, with **b-c**, Kaplan-Meier curves for **b**, Metastasis-free survival and **c**, Overall analysis for the selected cohort.

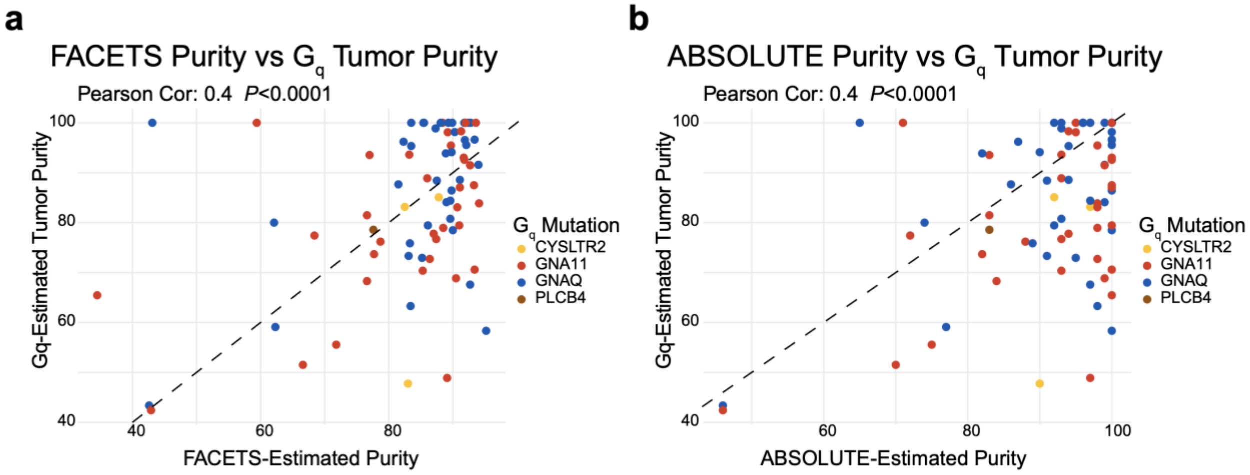


**Supplementary Figure 7.** Validation of tumor purity estimation by $G_q$ mutation VAF by comparison to copy number-based methodologies reported for UM within the TCGA cohort. Correlation plots of $G_q$ mutation-based tumor purity with previously reported purity estimates calculated with **a**, FACETS and **b**, ABSOLUTE algorithms. Plots include all UM cases with a detectable $G_q$ mutation (n=78). The dashed line indicates the identity line (y=x), while dot color indicates the representative $G_q$ mutation. Significance determined by two-tailed t-test. Pearson cor., Pearson correlation; FACETS, Fraction and Allelic Copy number Estimation from Tumor/normal Sequencing.
